# Risk factors for extended-spectrum beta-lactamase (ESBL) producing *E. coli* carriage among children in a food animal producing region of Quito, Ecuador

**DOI:** 10.1101/2022.11.14.22282279

**Authors:** Heather K. Amato, Fernanda Loayza, Liseth Salinas, Diana Paredes, Daniela Garcia, Soledad Sarzosa, Carlos Saraiva-Garcia, Timothy J. Johnson, Amy J. Pickering, Lee W. Riley, Gabriel Trueba, Jay P. Graham

**Author notes:** Recently deceased (Lee W. Riley).

## Abstract

**Background:** The spread of antibiotic-resistant bacteria may be driven by human-animal-environment interactions, especially in regions with limited restrictions on antibiotic use, widespread food animal production, and free-roaming domestic animals. In this study, we aimed to identify risk factors related to domestic animals, backyard food animals, and commercial food animal production in Ecuadorian communities.

**Methods & Findings:** We conducted a repeated-measures study from 2018-2021 in 7 semi-rural parishes of Quito, Ecuador to identify determinants of third-generation cephalosporin-resistant *E. coli* (3GCR-EC) and extended-spectrum beta-lactamase *E. coli* (ESBL-EC) in children and domestic animals. We used multivariable log-binomial regression models to estimate relative risks (RR) of 3GCR-EC and ESBL-EC carriage. We collected 1,699 child fecal samples from 600 households and 1,871 animal fecal samples from 376 of the same households. Risk factors for 3GCR-EC included living within 5 km of more than 5 commercial food animal operations (RR: 1.36; 95% Confidence Interval: 1.16, 1.59), household pig ownership (1.23; 1.02, 1.48), child pet contact (1.23; 1.09, 1.39), and rarely/never washing hands after contact with animals (1.15; 0.98, 1.34). Risk factors for ESBL-EC were dog ownership (1.43; 1.00, 2.04), child pet contact (1.54; 1.10, 2.16), placing animal feces on household land/crops (1.63; 1.09, 2.46), and combined exposures to both household food animals and commercial food animal operation drainage paths (1.80; 0.94, 3.45).

**Conclusions:** Policies and interventions that improve the safety of animal waste management in communities and in commercial food animal production operations may be necessary to curb the spread of resistant bacteria.

## Introduction

Widespread environmental contamination from food animal production is increasingly recognized as an important contributor to the global antibiotic resistance crisis. Globally, large quantities of clinically important antibiotics are administered to food animals (poultry and livestock raised for meat and dairy products) to promote growth and prevent infection (1). Antibiotic-resistant bacteria, including extended-spectrum beta-lactamase producing Enterobacterales (ESBL-E), found in humans have been linked to food animals (2). ESBL-E – deemed a serious threat to global public health by the World Health Organization and the U.S. Centers for Disease Control and Prevention – confer resistance to a broad spectrum of beta-lactam antibiotics including penicillins and cephalosporins, the most commonly used treatments for bacterial infections (3,4). With the rapid growth of the food animal production industry in low- and middle-income countries (LMICs), antibiotic use in these settings is projected to increase by upwards of 200% from 2010 to 2030, fueling the emergence and selection of these multidrug-resistant bacteria (1). Estimating the risks of ESBL-E colonization in communities with exposures to food animal production is a crucial step towards developing strategies to combat the global spread of antibiotic-resistant bacteria.

An estimated 1.27 million deaths in 2019 were attributable to bacterial antibiotic-resistant infections, 89% of which occurred in LMICs (5). Third-generation cephalosporins, a group of beta-lactam antibiotics, are used in hospitals to treat life-threatening infections. With limited treatment options, third-generation cephalosporin-resistant and ESBL-producing enterobacteriaceae infections result in longer and more costly hospital stays, increased severity of illness, and increased risk of mortality (6–9). Even in healthy individuals, asymptomatic carriage of ESBL-E in commensal gut bacteria may still pose a threat to health; ESBLs are frequently encoded by plasmids which facilitate horizontal transfer of resistance genes, allowing commensal bacteria to share ESBL-encoded genes with pathogenic bacteria (10–13). Horizontal gene transfer rapidly propagates phenotypic resistance among diverse bacteria in animals, the environment, and humans (10,14). Globally, the prevalence of ESBL-E is increasing; as of 2018, an estimated 20% of healthy individuals harbor ESBL-producing *Escherichia coli* in their guts (15).

In upper middle-income countries like Ecuador, both commercial and small-scale or “backyard” food animal production are increasing as population growth and increasing wealth drive consumption of animal products (16). Antibiotics are largely unregulated in Ecuador and other LMICs; medically important antibiotics are routinely used in large-scale, commercial food animal operations at subtherapeutic doses to increase feed conversion efficiency (17,18). Antibiotics are available over-the-counter without need for veterinarian prescription at local animal feed stores, and are also used in small-scale food animal production for growth promotion and disease prevention (17). In Ecuador, an estimated 84% of rural households and 29% of urban households own livestock, and small-scale poultry farmers have reported regularly administering a range of six different classes of antibiotics (19,20).

Contact with animal waste is elevated in LMICs, increasing the potential for exposure to ESBL-E and other antibiotic-resistant bacteria. Domestic animals and backyard food animals commonly defecate in the household environment resulting in fecal pathogen contamination (21,22). Young children are frequently exposed to high doses of poultry, livestock, and domestic animal feces through the consumption of soil and hand-to-mouth behaviors (23,24). Domestic animals and fecal contamination of household soil, food, and drinking water have been identified as sources of resistant bacteria in Nigeria, Peru, Brazil, India, and elsewhere (25–28). Exposure to animal feces can also increase the risk of diarrhea (29–31). Exposure and risk assessments and epidemiological studies in LMICs have focused on exposures to feces or fecal pathogens, broadly; few epidemiological studies have assessed exposures and risk factors for antibiotic-resistant and ESBL-E infections among children in LMICs (32–35).

Given the anticipated growth in unrestricted antibiotic use for food animals in LMICs, there is an urgent need to quantify the risks of antibiotic-resistant and ESBL-E infections among children exposed to small-scale and/or commercial food animal production in these settings. This study aimed to estimate the risk of cephalosporin-resistant and ESBL-producing *E. coli* carriage among children with varying degrees of exposure to small-scale and/or commercial food animal production in semi-rural parishes of Quito, Ecuador. We hypothesized *a priori* that (a) household-level exposures to small-scale food animal production are associated with an increased risk of third-generation cephalosporin-resistant *E. coli* (3GCR-EC) and ESBL-producing *E. coli* (ESBL-EC); (b) exposures to commercial food animal production operations in the community are associated with an increased risk of 3GCR-EC and ESBL-EC; and (c) combined exposures to both small-scale and commercial food animal production are associated with a greater increased risk of 3GCR-EC and ESBL-EC in children than small-scale or commercial food animal exposures, alone.

## Methods

This repeated measures observational study was carried out in semi-rural communities east of Quito, Ecuador by researchers at the Instituto de Microbiología at the Universidad San Francisco de Quito (USFQ) and the University of California, Berkeley School of Public Health. The study area was approximately 320 km^2^ and included commercial food animal operations and backyard food animal production. The study aimed to enroll 360 households through stratified random sampling across seven semi-rural parishes. Households were enrolled if they met the following inclusion criteria at the time of enrollment: (1) a primary child caretaker who was over 18 years of age was present, (2) a child between the ages of 6 months and 5 years old was present in the household; and (3) informed consent was provided by the caretaker to participate in the study. If there was more than one child at a given household, the youngest child was selected for participation. Households were visited up to five times between August 2018 and August 2021 by trained field staff who conducted household surveys and collected household GPS coordinates and biological samples at each visit. The study was approved by the Office for Protection of Human Subjects (OPHS) at the University of California, Berkeley (IRB# 2019-02-11803) and by the Bioethics Committee at the Universidad San Francisco de Quito (#2017-178M), and the Ecuadorian Health Ministry (#MSPCURI000243-3).

### Exposure Assessment

The primary exposures of interest were exposures to commercial food animal production in the community and household-level exposures to small-scale food animal production. Exposures and other household characteristics and practices were assessed at each household visit to capture time-varying exposures. Exposure to commercial food animal production was assessed in three ways: (1) distance to the nearest commercial food animal production facility; (2) density of commercial food animal production facilities; and (3) proximity to drainage paths of commercial food animal production facilities. Commercial poultry production facilities – vertically integrated operations that were marked by long barns with a metal roof and typically held approximately 20,000 birds or more – were located using satellite imagery in Google Maps and confirmed as active operations through site visits and ground-truthing. Other types of food animal production operations were identified through local knowledge and ground-truthing.

For the first measure of commercial food animal production exposure, euclidean distance between each household and the nearest active commercial food animal operation was measured at each time point. For the second measure of exposure, the density of commercial food animal operations was assessed by summing the number of operations within a five-kilometer buffer of each household at each time point. We used the *sf* package in R to create these first two exposure variables (36,37). While proximity may serve as a proxy for the likelihood of environmental contamination from nearby commercial operations, density may serve as a proxy for the extent of environmental contamination from nearby operations. Density of commercial poultry operations has been associated with cephalosporin-resistant *E. coli* in nearby stream water and sediment (38). Finally, for the third exposure measure, drainage paths from each commercial food animal operation were identified using ArcGIS Online Trace Downstream tool, which uses a digital elevation model to identify downstream flow paths from elevation surfaces, drainage directions, river networks, and watershed boundaries obtained from the HydroSHEDS 90m database (39). We then created buffers around drainage paths, which ranged from 2.9-3.1 km in length, and spatially joined study households to buffer layers in QGIS to identify which households were within 100 or 500 meters of commercial operation drainage paths (40) (Figure 1). Proximity to these drainage paths may capture exposures to antibiotic-resistant bacteria transported from commercial food animal operations through waterways, even when households are located further from the food animal operation, itself.

**Figure 1.**
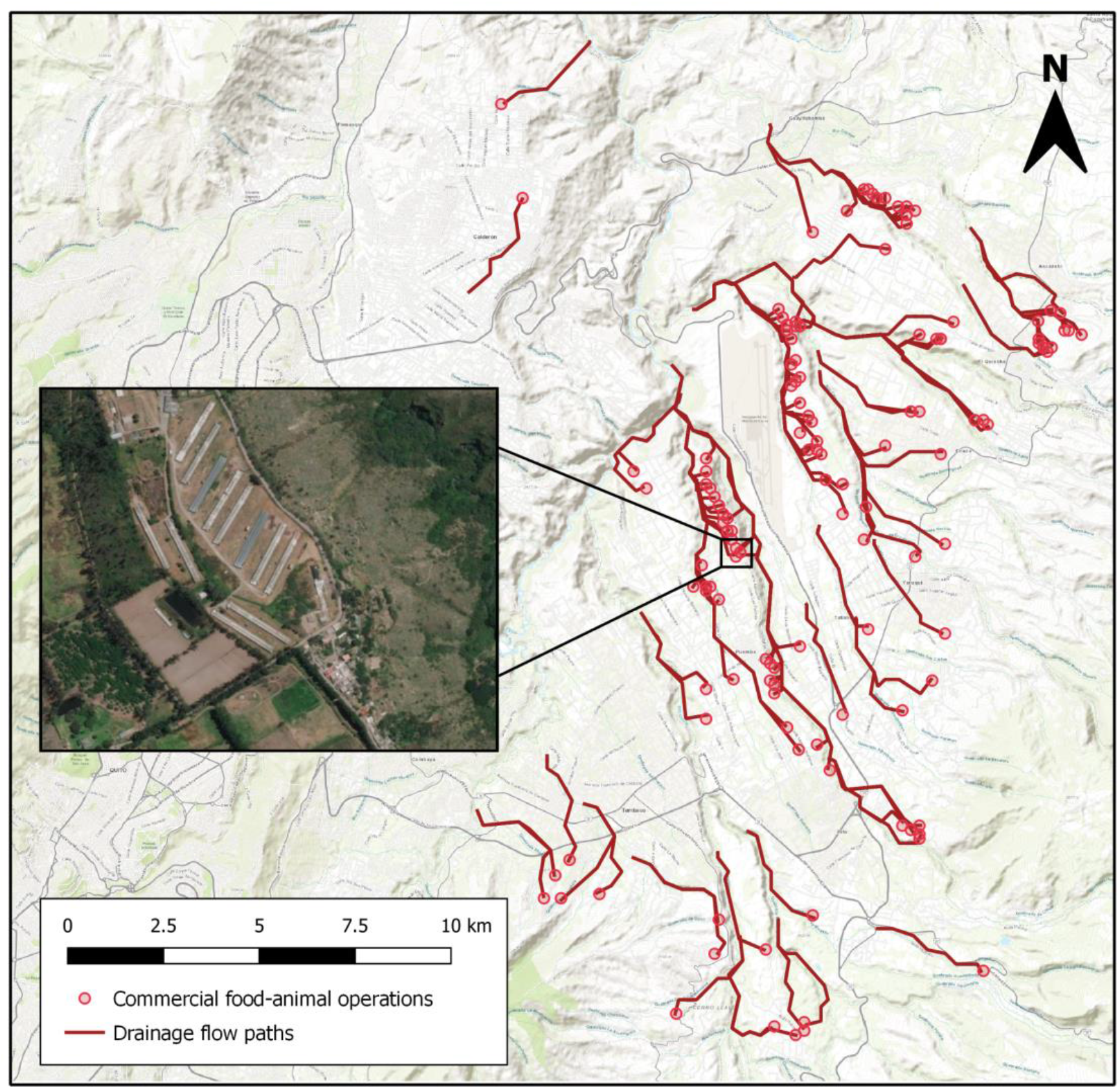
Map of active commercial food animal production operations (*n*=130) and their drainage flow paths in the study area, east of Quito, Ecuador.

Household-level exposure to small-scale food animal production was defined as household ownership of any food animals (i.e., chickens, pigs, cattle, sheep, goats, ducks, guinea pigs, rabbits, or quail). Household surveys captured information on caregiver-reported household ownership of food animals and other domestic animals, as well as household characteristics, caregiver and child demographics, health status, and other potential risk factors. Surveys were created using Open Data Kit (ODK) Build and trained enumerators used ODK Collect on Android devices for mobile data collection (41). Encrypted survey forms were sent to ODK Aggregate on a secure server at USFQ upon completion and were subsequently downloaded for de-identification and analysis.

### Outcome Assessment

A single stool sample from the youngest eligible child per household was collected at each visit to determine enteric carriage of third-generation cephalosporin resistant *E. coli* (3GCR-EC) and ESBL production based on phenotypic testing, described below. Caregivers were given supplies to collect a child stool sample, which were double-bagged and stored in a fridge or on ice (4°C), and the enumerator returned to collect the sample the next day. In the case that a child had not defecated before the enumerator returned, the enumerator came back to the household the following day to collect the sample. If animals were present at the household at the time of data collection, a single stool sample per animal species was collected from the environment where the animals defecate. Fecal samples were placed in sterile containers and stored on ice (4°C) during transportation to the microbiology lab at USFQ. Samples were processed at USFQ within five hours of collection to identify 3GCR-EC and conduct antibiotic susceptibility testing.

### Microbiological Methods

To screen for 3GCR-EC and improve sensitivity for detecting ESBL production among both dominant and non-dominant strains of *E. coli*, fecal samples were plated on MacConkey agar (Difco, Sparks, Maryland) with 2 mg/L of ceftriaxone and incubated for 18 hours at 37 ºC (42). Up to five isolates phenotypically matching *E. coli* were selected from each fecal sample and preserved at −80 ºC in Trypticase Soy Broth medium (Difco, Sparks, MD) with 20% glycerol. 3GCR-EC isolates for each fecal sample were thawed and regrown on MacConkey agar at 37 °C for 18-24 h for evaluation of antibiotic susceptibility by the disk diffusion method (Kirby Bauer test) on Mueller–Hinton agar (Difco, Sparks, Maryland). To confirm presumptive *E. coli* isolates, colonies were inoculated onto Chromocult® coliform agar (Merck, Darmstadt, Germany).

### Antibiotic Susceptibility Testing

Antibiotic susceptibility testing of all 3GCR-EC isolates was conducted for 10 antibiotics: ampicillin (AM; 10 μg), ceftazidime (CAZ; 30 μg), ciprofloxacin (CIP; 5 μg), cefotaxime (CTX; 30 μg), cefazolin (CZ; 30 μg), cefepime (FEP; 30 μg), gentamicin (GM; 10 μg), imipenem (IPM; 10 μg), trimethoprim/sulfamethoxazole (SXT; 1.25 per 23.75 μg), and tetracycline (TE; 30 μg). Isolates were identified as either susceptible or resistant to each antibiotic according to the resistance or susceptibility interpretation criteria from Clinical and Laboratory Standards Institute (CLSI) guidelines (43). *E. coli* ATCC 25922 was used as the quality control strain. Multidrug resistance (resistant to 3 or more classes) was determined based on the number of macro-classes to which each isolate was resistant. Macro-classes were defined as cephalosporin/beta-lactamase inhibitors, penicillins, aminoglycosides, carbapenems, fluoroquinolones, tetracyclines, and folate pathway inhibitors.

For phenotypic confirmation of ESBL production, the combination disk diffusion test was used with CAZ and CAZ/CLA (ceftazidime with clavulanic acid) as outlined in the CLSI guidelines (43). In the first four cycles of data collection, up to five *E. coli* isolates per sample were selected and preserved for analysis. 3GCR-EC isolates from the same fecal sample with identical phenotypic resistance profiles were considered duplicates and were de-duplicated prior to analyses. Due to limited laboratory resources after the COVID-19 pandemic began in 2020 and the high rate of clonal relationships between *E. coli* isolated from the same sample (based on preliminary sequencing), only one *E. coli* isolate per sample was selected and preserved for analysis during the fifth cycle of data collection.

### DNA Sequencing and Analysis

Genomic DNA was extracted from the isolates using Wizard® Genomic DNA Purification (Promega) kits and QIAGEN© DNEasy Blood & Tissue Kits according to the manufacturer’s instructions. Whole-genome sequencing was carried out at the University of Minnesota. In brief, we sequenced whole-genome *E. coli* isolates using either Illumina MiSeq or NovaSeq with Nextera XT libraries. Following sequencing, raw reads were quality-trimmed and adapter-trimmed using trimmomatic (44). Assemblies of reads was performed using SPAdes (Bankevich et al. 2012), then antibiotic resistance genes (ARGs) were identified using ABRicate (version 0.8.13) and a curated version of the ResFinder database (Zankari et al. 2012). We also performed in silico multilocus sequence typing (MLST) based on seven housekeeping genes (adk, fumC, gyrB, icd, mdh, purA, and recA), an additional eight housekeeping genes (dinB, icdA, pabB, polB, putP, trpA, trpB, and uidA), and core genome (cgMLST) using MLST 2.0 (45) and cgMLSTFinder 1.1 (46). Detailed methods are previously described (47).

### Statistical Analyses

Multivariable log-binomial regression models were used to estimate adjusted relative risks (RRs) for the associations between commercial or household food animal exposures and 3GCR-EC and ESBL-EC carriage, accounting for a potential interaction between household food animal ownership and each measure of commercial food animal exposures in three different models. Exposures were treated as binary (presence/absence of household food animals) and categorical variables (exposures to commercial food animal operations) using cut-points selected based on a combination of factors, such as the data distribution and policy relevance. The primary outcomes, 3GCR-EC and ESBL-EC carriage, were assessed at the isolate level and treated as binary. Each model controlled for pre-specified covariates (child age, child sex, child antibiotic use in the past 3 months, household asset score as a proxy for socioeconomic status, and caregiver education level) identified using a directed acyclic graph and existing literature (Supplemental Materials, Figure S1) (48).

We estimated the effects of each exposure individually and the combined effects of each measure of exposure to commercial food animals given household food animal ownership, comparing combined effects to a single referent group (unexposed to both commercial and household food animals). We also assessed effect measure modification by estimating the effect of each measure of exposure to commercial food animals among those with vs. without food animals (i.e., stratum-specific effects). Estimates and *P*-values from interaction models were used to determine multiplicative interaction based on an alpha level of < 0.10. In a sensitivity analysis, we reproduced this main analysis using only the first *E. coli* isolate per fecal sample to assess the potential for bias due to the change in number of isolates and probability of detecting the outcomes at each time point. Additional log-binomial regression models estimated associations between secondary risk factors of interest related to household animal ownership and animal contact. All regression models used generalized estimating equations with an exchangeable working correlation to obtain robust standard errors, adjusting for repeated measures and for the unbalanced data structure. Statistical analyses and visualizations were completed in R version 3.6.1 (37) using the *dplyr* (49), *tableone* (50), *ggplot2* (51), *geepack* (52), *multcomp* (53), and *car* (54) packages.

### Results

A total of 605 households across seven semi-rural parishes east of Quito, Ecuador were enrolled throughout the study period between July 2018 and September 2021. We enrolled 374 households in the initial cycle of data collection and recruited new households (using the same enrollment criteria) to enroll in subsequent data collection cycles to account for loss to follow-up. During the fourth cycle, data collection was halted in March 2020 due to lockdown restrictions during the SARS-CoV2 pandemic, resulting in significant loss to follow-up (39.7%, 151/380 lost to follow-up). However, in the fifth cycle during 2021, we re-enrolled 63.4% (241/380) of participants from cycle three and 78.8% (186/236) of participants from cycle four (Figure S2). Eleven (1.8%) of the 605 households enrolled in total were missing either exposure, outcome, or covariate data and were not included in statistical analyses.

A total of 1,739 child fecal samples were collected throughout the study period, from which 920 distinct 3GCR-EC colonies were isolated. Forty (2.3%) child fecal samples were missing corresponding survey data, resulting in a total of 910 3GCR-EC isolates from 1,699 child fecal samples across 600 households. We identified 1,060 3GCR-EC isolates from 1,871 animal fecal samples collected at 376 of the same households. After removing children with missing exposure, outcome, and covariate data, 904 3GCR-EC isolates from 1,677 child fecal samples from 594 households remained, resulting in a total of 1,940 observations in the final dataset for the primary statistical analysis.

### Household & Child Characteristics

In a majority of households, most primary caregivers had at least a high school or college level education, ranging from 67.1-79.5% across the data collection cycles (Table 1). The average age of children participating in the study was 1.8 years during the first cycle of data collection (Table 2). Access to drinking water and sanitation was high; 98.8% of households had a flush toilet (to sewer or septic tank), 92.2% had piped drinking water inside their home, and 98.6% had 24-hour access to drinking water (data not shown). Over 93% of households had household handwashing stations with the presence of both soap and water, confirmed by observation (Table 1). However, caregiver-reported child hand washing frequency suggested that most children rarely (44.4-69.6%) washed their hands after contact with animals (Table 2). Over the course of the entire study period, 36.7% of caregivers reported that their child played near animal feces in the last 3 weeks, 36.0% reported that their child had contact with livestock at least once per week in the last three months, and 67.9% reported child contact with pets at least once per week in the last 2021 during the SARS-CoV2 pandemic (Table 2). Similarly, treatment for infection in the last three months declined from 31.1% at the first cycle to 8.1% at the fifth cycle; child antibiotic use in the last three months also declined from 26.2% at cycle one to 5.9% at cycle five (Table 2).

**Table 1.** Characteristics of study households in semi-rural Quito, Ecuador at each cycle of data collection between July 2018 and September 2021.

|  | Data Collection Cycle |  |  |  |  |
| --- | --- | --- | --- | --- | --- |
|  | 1<br>*n* (%) | 2<br>*n* (%) | 3<br>*n* (%) | 4<br>*n* (%) | 5<br>*n* (%) |
| *Total* | 370 (100) | 358 (100) | 365 (100) | 225 (100) | 356 (100) |
| *Parish* |  |  |  |  |  |
| El Quinche | 20 (5.4) | 17 (4.7) | 30 (8.2) | 16 (7.1) | 26 (7.3) |
| Puembo | 20 (5.4) | 21 (5.9) | 37 (10.1) | 16 (7.1) | 23 (6.5) |
| Pifo | 86 (23.2) | 77 (21.5) | 75 (20.5) | 55 (24.4) | 75 (21.1) |
| Tababela | 12 (3.2) | 9 (2.5) | 8 (2.2) | 6 (2.7) | 13 (3.7) |
| Tumbaco | 23 (6.2) | 35 (9.8) | 24 (6.6) | 17 (7.6) | 22 (6.2) |
| Yaruqui | 146 (39.5) | 146 (40.8) | 141 (38.6) | 80 (35.6) | 141 (39.6) |
| Missing | 4 (1.1) | 0 (0.0) | 0 (0.0) | 0 (0.0) | 0 (0.0) |
| *Caregiver education level* |  |  |  |  |  |
| High school or college | 276 (74.6) | 263 (73.5) | 256 (70.1) | 151 (67.1) | 283 (79.5) |
| Elementary | 93 (25.1) | 95 (26.5) | 108 (29.6) | 68 (30.2) | 69 (19.4) |
| Missing | 1 (0.3) | 0 (0.0) | 1 (0.3) | 6 (2.7) | 4 (1.1) |
| *Household size (mean (SD))* | 4.5 (1.4) | 4.6 (1.5) | 4.6 (1.4) | 4.4 (1.2) | 4.4 (1.5) |
| *Household crowding\* (mean (SD))* | 2.6 (1.1) | 2.5 (1.0) | 2.5 (1.0) | 2.4 (0.9) | 2.2 (0.8) |
| *Household water treatment* |  |  |  |  |  |
| No treatment | 194 (52.4) | 170 (47.5) | 223 (61.1) | 138 (61.3) | 229 (64.3) |
| Boil | 159 (43.0) | 154 (43.0) | 119 (32.6) | 68 (30.2) | 109 (30.6) |
| Chlorinate or filter | 3 (0.8) | 5 (1.4) | 5 (1.3) | 3 (1.3) | 2 (0.6) |
| Other | 14 (3.8) | 29 (8.1) | 18 (4.9) | 16 (7.1) | 16 (4.5) |
| *Household handwashing station* |  |  |  |  |  |
| Soap & water | 354 (95.7) | 344 (96.1) | 347 (95.1) | 211 (93.8) | 340 (95.5) |
| Soap or water, only | 9 (2.4) | 14 (4.0) | 17 (4.7) | 14 (6.2) | 16 (4.5) |
| Neither | 6 (1.6) | 0 (0.0) | 1 (0.3) | 0 (0.0) | 0 (0.0) |
| Missing | 1 (0.3) | 0 (0.0) | 0 (0.0) | 0 (0.0) | 0 (0.0) |
| *Caregiver worked with animals in past 6 months\** | 105 (28.4) | 79 (22.1) | 80 (21.9) | 73 (32.4) | 113 (31.7) |
| *Household owns animals* | 233 (63.0) | 241 (67.3) | 244 (66.8) | 157 (69.8) | 236 (66.3) |
| Missing | 3 (0.8) | 1 (0.3) | 0 (0.0) | 0 (0.0) | 0 (0.0) |
| *Household owns food animals* | 110 (29.7) | 117 (32.7) | 107 (29.3) | 63 (28.0) | 92 (25.8) |
| Missing | 2 (0.5) | 1 (0.3) | 0 (0.0) | 0 (0.0) | 0 (0.0) |
| *Household food animals (mean (SD))* | 6.9 (19.3) | 5.7 (14.1) | 6.9 (19.1) | 9.2 (39.4) | 5.9 (21.3) |
| *Antibiotic use for household food animals* | 29 (26.4) | 22 (18.8) | 11 (10.3) | 6 (9.5) | 0 (0.0) |
| Missing | 7 (6.4) | 4 (3.4) | 1 (0.9) | 0 (0.0) | 9 (9.8) |
| *Distance to nearest commercial food animal operation* |  |  |  |  |  |
| ≥ 1.5 km | 155 (41.9) | 162 (45.3) | 154 (42.2) | 97 (43.1) | 170 (47.8) |
| 1-1.49 km | 90 (24.3) | 86 (24.0) | 87 (23.8) | 52 (23.1) | 70 (19.7) |
| 0.5-0.9 km | 60 (16.2) | 54 (15.1) | 73 (20.0) | 45 (20.0) | 71 (19.9) |
| < 0.5 km | 61 (16.5) | 56 (15.6) | 51 (14.0) | 31 (13.8) | 45 (12.6) |
| Missing | 4 (1.1) | 0 (0.0) | 0 (0.0) | 0 (0.0) | 0 (0.0) |
| *Commercial food animal operations within 5 km radius* |  |  |  |  |  |
| ≤ 5 | 115 (31.1) | 118 (33.0) | 113 (31.0) | 71 (31.6) | 203 (57.0) |
| 6-10 | 93 (25.1) | 89 (24.9) | 84 (23.0) | 52 (23.1) | 44 (12.4) |
| 11-20 | 88 (23.8) | 90 (25.1) | 76 (20.8) | 51 (22.7) | 47 (13.2) |
| > 20 | 70 (18.9) | 61 (17.0) | 92 (25.2) | 51 (22.7) | 62 (17.4) |
| Missing | 4 (1.1) | 0 (0.0) | 0 (0.0) | 0 (0.0) | 0 (0.0) |
| *Distance to drainage path of commercial food animal operation* |  |  |  |  |  |
| > 500 m | 154 (41.6) | 161 (45.0) | 150 (41.1) | 99 (44.0) | 167 (46.9) |
| 101-500 m | 158 (42.7) | 133 (37.2) | 162 (44.4) | 92 (40.9) | 138 (38.8) |
| ≤ 100 m | 60 (16.2) | 67 (18.7) | 59 (16.2) | 39 (17.3) | 56 (15.7) |
| Missing | 4 (1.1) | 0 (0.0) | 0 (0.0) | 0 (0.0) | 0 (0.0) |

**Table 2.** Characteristics and behaviors of children in study households at each cycle of data collection between July 2018 and September 2021.

|  | Data Collection Cycle |  |  |  |  |
| --- | --- | --- | --- | --- | --- |
|  | 1<br>*n* (%) | 2<br>*n* (%) | 3<br>*n* (%) | 4<br>*n* (%) | 5<br>*n* (%) |
| *Total* | 370 (100) | 358 (100) | 365 (100) | 225 (100) | 356 (100) |
| *Child age in years (mean (SD))* | 1.8 (1.3) | 2.0 (1.3) | 2.4 (1.4) | 3.0 (1.3) | 3.8 (1.6) |
| *Child sex* |  |  |  |  |  |
| Female | 171 (46.2) | 162 (45.3) | 154 (42.2) | 96 (42.7) | 158 (44.4) |
| Male | 199 (53.8) | 196 (54.7) | 211 (57.8) | 129 (57.3) | 198 (55.6) |
| *Child contact with livestock in last 3 months* |  |  |  |  |  |
| Never | 248 (67.0) | 254 (70.9) | 238 (65.2) | 126 (56.0) | 205 (57.6) |
| <1 time per week | 42 (11.4) | 26 (7.3) | 54 (14.8) | 14 (6.2) | 25 (7.0) |
| 1-2 times per week | 31 (8.4) | 32 (8.9) | 39 (10.7) | 50 (22.2) | 50 (14.0) |
| 3+ times per week | 49 (13.2) | 46 (12.8) | 34 (9.3) | 35 (15.6) | 75 (21.1) |
| Missing | 0 (0.0) | 0 (0.0) | 0 (0.0) | 0 (0.0) | 1 (0.3) |
| *Child contact with pets in last 3 months* |  |  |  |  |  |
| Never | 132 (35.7) | 122 (34.1) | 108 (29.6) | 64 (28.4) | 112 (31.5) |
| <1 time per week | 54 (14.6) | 34 (9.5) | 65 (17.8) | 12 (5.3) | 23 (6.5) |
| 1-2 times per week | 52 (14.1) | 50 (14.0) | 73 (20.0) | 59 (26.2) | 50 (14.0) |
| 3+ times per week | 132 (35.7) | 152 (42.5) | 119 (32.6) | 90 (40.0) | 171 (48.0) |
| *Child played near animal feces in last 3 weeks* | 115 (31.1) | 99 (27.7) | 148 (40.5) | 77 (34.2) | 175 (49.2) |
| Missing | 2 (0.5) | 0 (0.0) | 0 (0.0) | 0 (0.0) | 3 (0.8) |
| *Child handwashing after contact with animals* |  |  |  |  |  |
| Never | 43 (11.6) | 13 (3.6) | 4 (1.1) | 3 (1.3) | 3 (0.8) |
| Rarely | 185 (50.0) | 249 (69.6) | 245 (67.1) | 129 (57.3) | 158 (44.4) |
| Sometimes | 83 (22.4) | 68 (19.0) | 109 (29.9) | 87 (38.7) | 159 (44.7) |
| Always | 18 (4.9) | 9 (2.5) | 7 (1.9) | 6 (2.7) | 36 (10.1) |
| Don't know/doesn't apply | 41 (11.1) | 19 (5.3) | 0 (0.0) | 0 (0.0) | 0 (0.0) |
| *Child had diarrhea in last 7 days* | 75 (20.3) | 77 (21.5) | 43 (11.8) | 36 (16.0) | 15 (4.2) |
| Missing | 2 (0.5) | 1 (0.3) | 1 (0.3) | 0 (0.0) | 2 (0.6) |
| *Child treated for infection in last 3 months* | 115 (31.1) | 98 (27.4) | 84 (23.0) | 54 (24.0) | 29 (8.1) |
| Missing | 3 (0.8) | 1 (0.3) | 0 (0.0) | 0 (0.0) | 0 (0.0) |
| *Child took antibiotics in last 3 months* | 97 (26.2) | 67 (18.7) | 63 (17.3) | 26 (11.6) | 21 (5.9) |
| Missing | 0 (0.0) | 2 (0.6) | 0 (0.0) | 0 (0.0) | 0 (0.0) |

### Food Animal Production & Domestic Animals

We identified 130 active commercial food animal operations in our study site including 122 poultry facilities, five hog facilities, two horse facilities, and one milk production facility. Across the seven parishes in the study site, four parishes (Tababela, Tumbaco, Checa (Chilpa), and Pifo) had low-intensity commercial production with fewer than 10 commercial food animal operations, each. Three parishes (El Quinche, Puembo, Yaruqui) had high-intensity production, with more than 30 commercial food animal operations, each. More than half of study households were located < 1.5 km from a commercial food animal operation (55.7%) or were within 500 m of a commercial operation’s drainage flow path (56.1%) throughout the study period. During the first four cycles of data collection, at least 67% of households were within 5 km of 6 or more commercial food animal operations (Table 1).

Throughout the study period, 66.4% of caregivers reported owning any type of animal, with 29.2% owning food animals and an average of 5.7-9.2 food animals at each data collection cycle (Table 1). Among households that owned food animals, reported antibiotic use in food animals was low, declining from 26.4% at the beginning of the study period to 0% at the end of the study (Table 1). The primary food animals owned were chickens. Thirty-two percent of households owned backyard chickens, 17% owned guinea pigs, 11% owned pigs, 7% owned cattle, and 3% owned goats or sheep. The average flock size for backyard chickens was 13.7 (Standard Deviation (SD): 29.9; range: 1-500). There were an average of 16 guinea pigs (SD: 19.6; range: 1-200), 4.3 pigs (SD: 6.6; range: 1-50), 2.8 cows (SD: 2.6; range: 1-20), 2.9 goats (SD: 3.2; range: 1-10), and 4.0 sheep (SD: 6.3; range: 1-30). Eighty percent of households owned dogs, with an average of 2.3 dogs per household (SD: 1.6; range: 1-12). Less than one third of caregivers reported working with animals (including live animals, animal feces, or meat processing) in the last six months during the study (Table 1).

### Characterization of 3GCR-EC Isolates

Sixty-one percent of children (n=365/600) were carriers of 3GCR-EC and 25% (n=149/600) were carriers of ESBL-EC at least once throughout the study period. 3GCR-EC were detected in 38% of child samples (n=652/1,699) and 51% of animal samples (n=959/1,871) from matched households. Of 910 3GCR-EC isolated from child samples, 36% were resistant to fourth-generation cephalosporin, cefepime, in phenotypic susceptibility testing (Table S2). Ninety-nine percent of child 3GCR-EC isolates were resistant to ampicillin, 64% were resistant to sulfamethoxazole/trimethoprim, 46% percent were resistant to ciprofloxacin, 16% were resistant to gentamicin, and <1% were resistant to imipenem (carbapenem) (Table S2). Eighty-six percent of child 3GCR-EC isolates were multidrug-resistant (3 or more classes), 37% were extensively drug-resistant (5 or more classes), and 22% percent were phenotypic ESBL producers (Table S2). Phenotypic resistance to antibiotics among 3GCR-EC isolates from animal fecal samples are described in Supplemental Materials (Table S3).

We analyzed whole-genome sequencing data for a subset of 571 3GCR-EC isolated from child fecal samples across 365 households. The most prevalent sequence type (ST) among these isolates was ST 10 (7%), while other clinically important STs like ST 131 and ST 117 accounted for about 3% of sequenced isolates (Table S4). 3GCR-EC isolates had a mean of 9.7 total antibiotic resistance genes (ARGs) (SD: 4.3), and 1.7 beta-lactamase (*bla*) genes (SD: 1.5). Overall, the proportion of 3GCR-EC isolates with *bla* genes was 65%, 52%, 8%, 5%, and 5% for *bla*_CTX-M_-encoding, *bla*_TEM_, *bla*_CMY_, *bla*_OXA_, and *bla*_SHV_ genes, respectively. Ten (2%) 3GCR-EC isolates had an *mcr-1* gene, indicating resistance to the last-line antibiotic colistin. Eighty-two percent (n=119/145) of phenotypic ESBL-EC and 77% (n=330/426) of non-ESBL-producing 3GCR-EC carried at least one *bla* gene. Among phenotypic ESBL-EC, the most prevalent *bla* genes were *bla*_CTX-M-55_, *bla*_TEM-141_, *bla*_TEM-1B_, *bla*_CTX-M-15_, and *bla*_OXA-1_ (Table S5, Figure S3). The most prevalent *bla* genes among 3GCR-EC that were not phenotypic ESBL-producers were *bla*_TEM-141_, *bla*_CTX-M-55_, *bla*_TEM-1B_, *bla*_CTX-M-65_, and *bla*_CTX-M-15_ (Table S5, Figure S3). Among the most prevalent CTX-M-encoding genes, the most prevalent genes in Yaruqui and Puembo - parishes with high-intensity commercial food animal production - were *bla*_CTX-M-55_, *bla*_CTX-M-65_, and *bla*_CTX-M-15_ (Figure 2, Table S8). El Quinche - another parish with high-intensity commercial production - had a higher prevalence of *bla*_CTX-M-8_ and *bla*_CTX-M-14,_ in addition to *bla*_CTX-M-55_, *bla*_CTX-M-65_, and *bla*_CTX-M-15_, compared to other parishes (Figure 2, Table S8). Tumbaco, Pifo, and Checa (Chilpa) - parishes with lower-intensity commercial food animal production - had a higher prevalence of *bla*_CTX-M-3_, in addition to *bla*_CTX-M-55_, *bla*_CTX-M-65_, and *bla*_CTX-M-15_, though *bla*_CTX-M-3_ was also detected in Yaruqui (Figure 2, Table S8).

**Figure 2.**
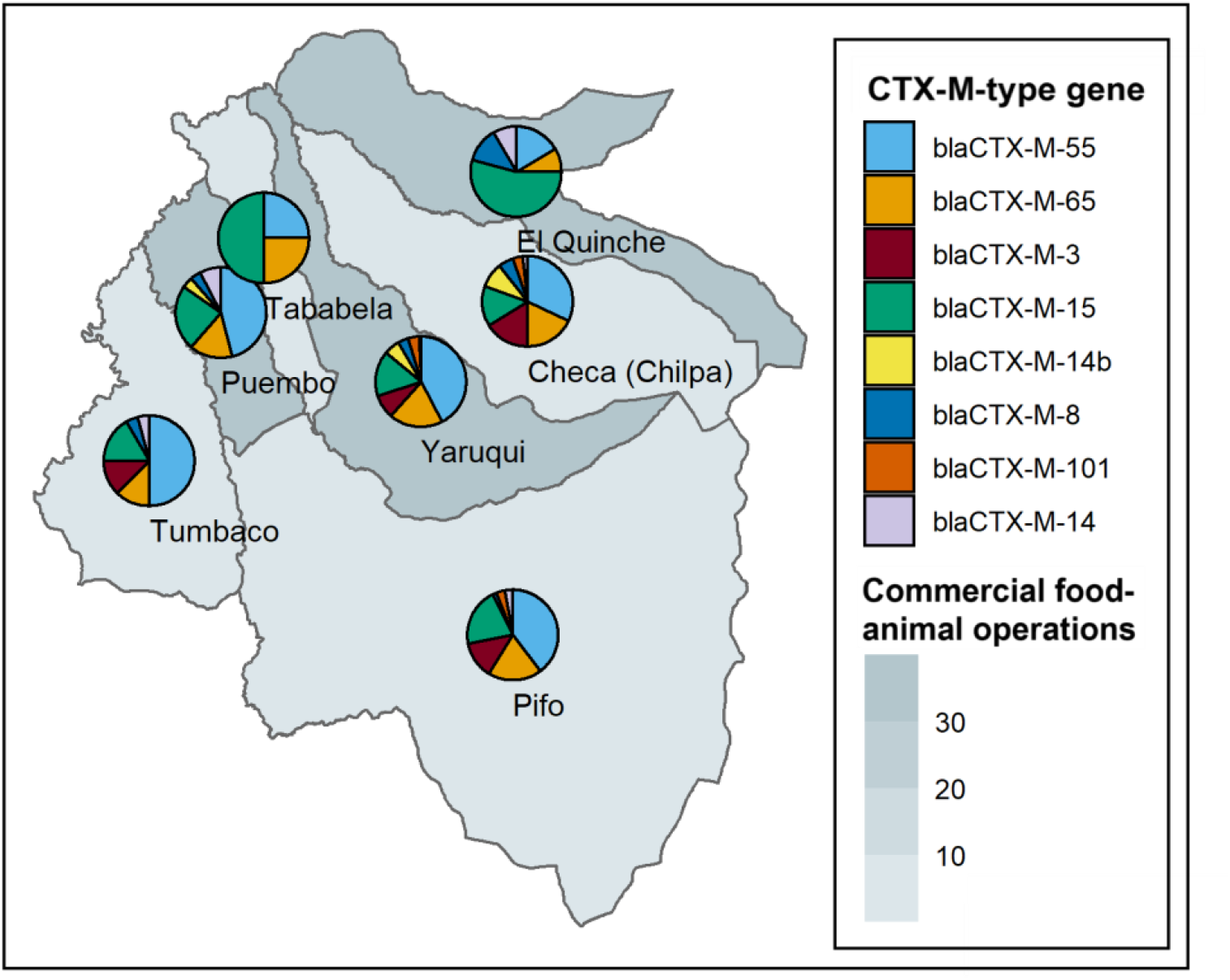
Distribution of CTX-M-type genes from a subset of 571 third-generation cephalosporin-resistant *E. coli* isolated from children across seven parishes east of Quito, Ecuador. Pie chart sections represent the proportion of isolates with a CTX-M-type gene detected, among those listed in the plot legend. Shaded areas represent parishes, where darker shading indicates a higher number of commercial food animal operations in a given parish.

### Risk Factors for 3GCR-EC

Children with > 5 commercial food animal operations in a 5-km radius of their household had 1.36 times the risk (95% Confidence Interval (CI): 1.16, 1.59) of 3GCR-EC carriage than those with ≤ 5 operations within 5 km (Table 3, Figure 3). However, among those with household food animals, there was no effect of commercial operation density on 3GCR-EC carriage (RR: 1.05; 95% CI: 0.83, 1.33) (Table 3, Figure 3). When controlling for the number of commercial food animal operations within 5 km, household food animal ownership was marginally associated with an increased risk of 3GCR-EC carriage among children (RR: 1.22; 95% CI: 0.95, 1.55) (Table 3, Figure 3). While the effect of commercial operation density was modified by household food animal ownership, there was not a significant excess risk of being exposed to both household food animals and >5 commercial operations (Relative Excess Risk due to Interaction (RERI): −0.29, 95% CI: −0.64, 0.06) (Table 3, Figure 3). Distance to the nearest commercial food animal operation was not significantly associated with the risk of 3GCR-EC carriage among households without food animals (RR: 1.09; 95% CI: 0.95, 1.27) (Table 3, Figure 3). However, the combination of owning household food animals and living <1.5 km from the nearest commercial operation was associated with an increased risk in 3GCR-EC carriage (RR: 1.15; 95% CI: 0.97, 1.33) compared to those without food animals who lived further from commercial food animal operations (Table 3, Figure 3). Proximity to a drainage flow path from a commercial food animal operation was not associated with 3GCR-EC carriage, regardless of household food animal ownership (Table 3, Figure 3). These results were largely robust to sensitivity analyses, which included only one 3GCR-EC isolate per sample (Table S9).

**Table 3.**
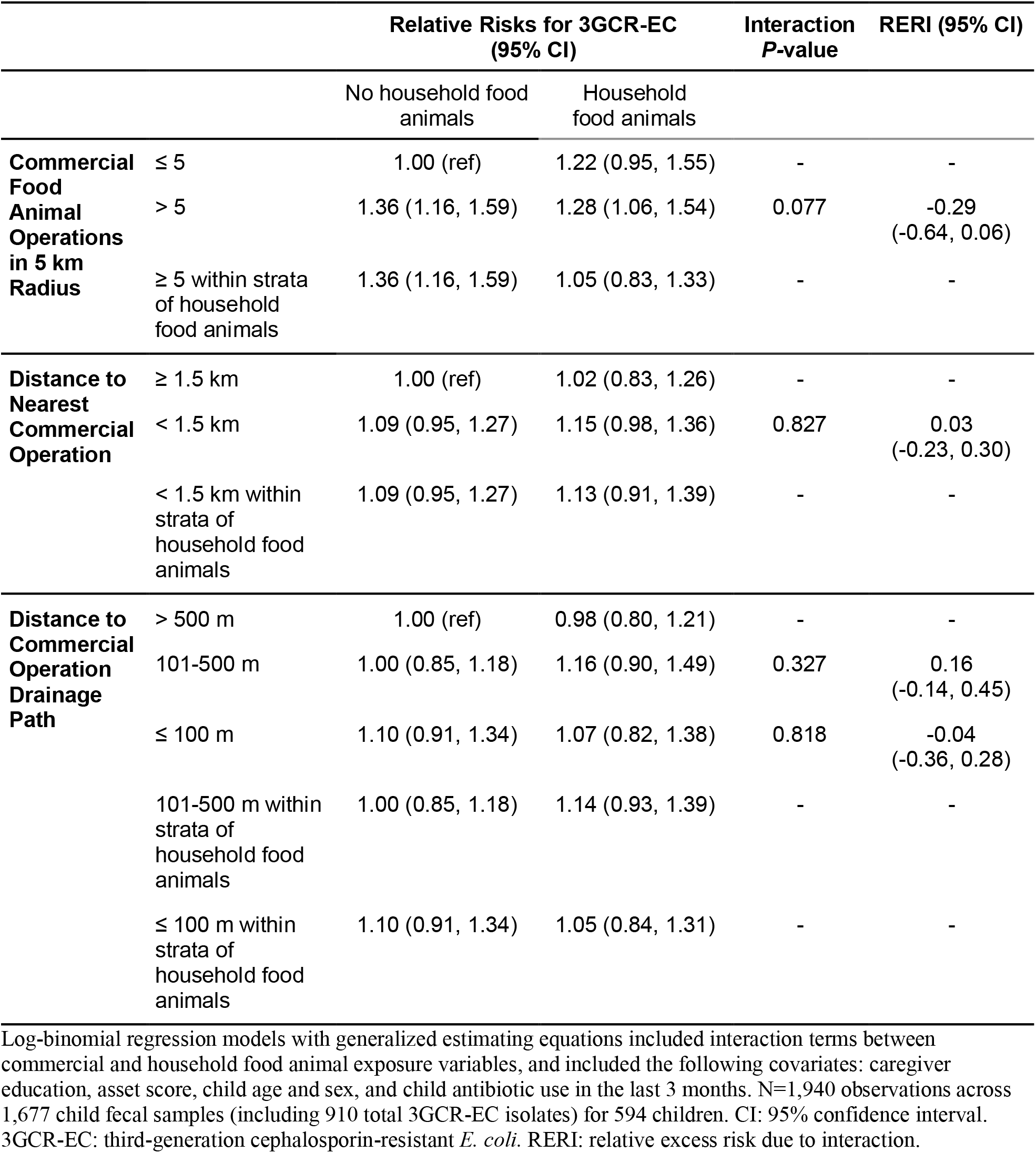
Relative risks of third-generation cephalosporin-resistant *E. coli* (3GCR-EC) carriage among children given exposures to commercial food animal production and effect measure modification by household food animal ownership.

**Figure 3.**
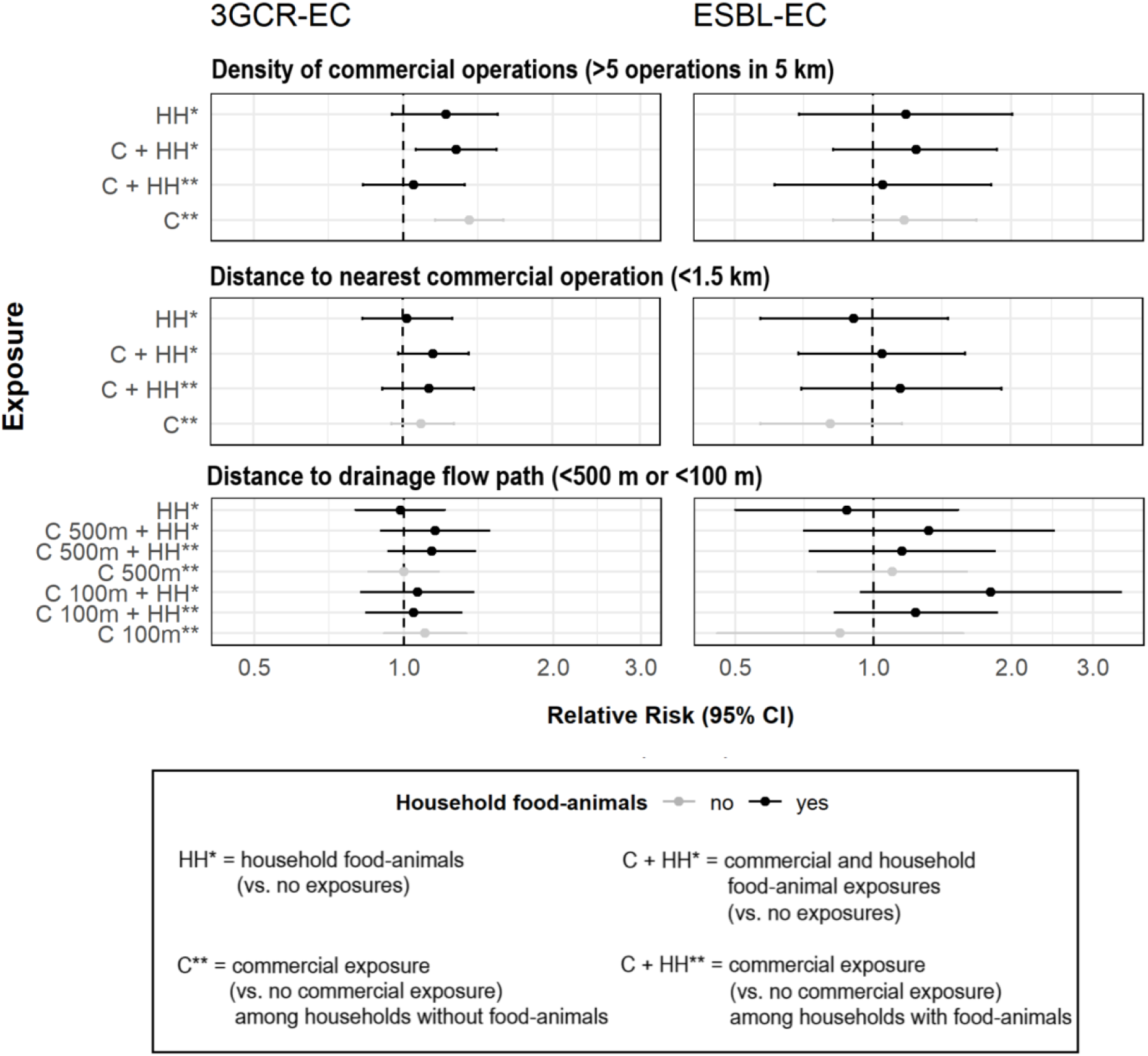
Interaction and stratum-specific effects of combined exposures to household and commercial food animal production on third-generation cephalosporin-resistant *E. coli* (3GCR-EC) and extended-spectrum beta-lactamase producing *E. coli* (ESBL-EC) carriage in children (N=1,940).

Other risk factors for 3GCR-EC among children included child pet contact in the last three months (RR: 1.23; 95% CI: 1.09, 1.39), pig ownership (RR: 1.23; 95% CI: 1.02, 1.48), chicken ownership (RR: 1.10; 95% CI: 0.98, 1.24), and rarely/never washing hands after contact with animals (RR: 1.15; 95% CI: 0.98, 1.34) (Figure 4, Table S10). Potential risk factors that were marginally associated with 3GCR-EC carriage included household animals being treated with antibiotics (RR: 1.15; 95% CI: 0.95, 1.39), household livestock/poultry drinking irrigation water (RR: 1.21; 95% CI: 0.94, 1.56), and the presence of ESBL-EC+ animal feces in the yard (RR: 1.11; 95% CI: 0.95, 1.30) (Figure 4, Table S10).

**Figure 4.**
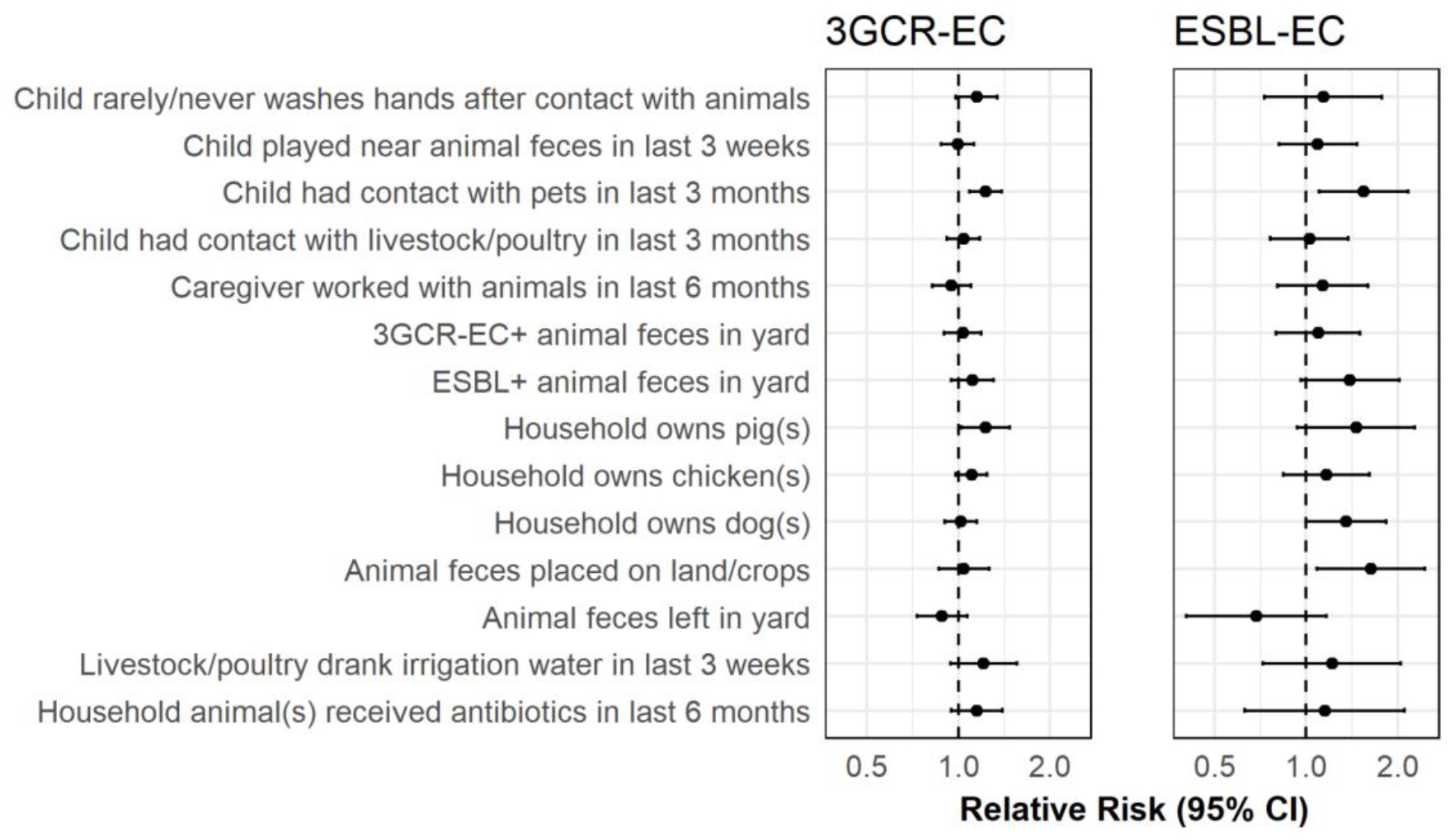
Risk factors for third-generation cephalosporin *E. coli* (3GCR-EC) and extended-spectrum beta-lactamase producing *E. coli* (ESBL-EC) carriage among children in semi-rural parishes of Quito, Ecuador (corresponding data in Table S7).

### Risk Factors for ESBL-EC

We did not detect significant associations or interactions between the density of or proximity to commercial food animal operations, household food animal ownership, and ESBL-EC carriage. Individually, household food animal ownership and increased proximity to a drainage path were not significantly associated with an increased risk of ESBL-EC carriage among children, but the effect sizes were larger for combined and increasing intensity of exposures. When controlling for household food animal ownership, the RR for ESBL-EC was 1.10 (95% CI: 0.76, 1.60) for those living 101-500 m from a drainage path compared to those > 500 m from a drainage path. The RR increased to 1.32 (95% CI: 0.71, 2.46) for those living 101-500 m from a drainage path and with household food animals compared to those > 500 m from a drainage path and without household food animals (Table 4, Figure 3). The RR increased further to 1.80 (95% CI: 0.94, 3.45) for those living within 100 m of a drainage path and with household food animals compared to those > 500 m from a drainage path and without household food animals; this interaction was near-significant with an interaction *P*-value of 0.1080 (RERI: 0.85; 95% CI: −0.10, 1.81) (Table 4, Figure 3). These findings were also robust to sensitivity analyses (Table S9).

**Table 4.** Relative risks of extended-spectrum beta-lactamase producing *E. coli* (ESBL-EC) carriage among children given exposures to commercial food animal production and effect measure modification by household food animal ownership.

|  |  | Relative Risks for ESBL-EC<br>(95% CI) |  | Interaction<br>*P*-value | RERI (95% CI) |
| --- | --- | --- | --- | --- | --- |
|  |  | No household<br>food animals | Household food<br>animals |  |  |
| **Commercial Food Animal Operations in 5 km Radius** | < 5 | 1.00 (ref) | 1.18 (0.69, 2.01) | - | - |
|  | ≥ 5 | 1.17 (0.82, 1.68) | 1.24 (0.82, 1.86) | 0.737 | -0.12<br>(-0.89, 0.66) |
|  | ≥ 5 within strata of household food animals | 1.17 (0.82, 1.68) | 1.05 (0.61, 1.81) | - | - |
| **Distance to Nearest Commercial Operation** | ≥ 1.5 km | 1.00 (ref) | 0.91 (0.57, 1.46) | - | - |
|  | < 1.5 km | 0.81 (0.57, 1.16) | 1.05 (0.69, 1.59) | 0.268 | 0.32<br>(-0.23, 0.88) |
|  | < 1.5 km within strata of household food animals | 0.81 (0.57, 1.16) | 1.15 (0.70, 1.91) | - | - |
| **Distance to Commercial Operation Drainage Path** | > 500 m | 1.00 (ref) | 0.88 (0.50, 1.53) | - | - |
|  | 101-500 m | 1.10 (0.76, 1.60) | 1.32 (0.71, 2.46) | 0.619 | 0.18<br>(-0.53, 0.89) |
|  | ≤ 100 m | 0.85 (0.46, 1.57) | 1.80 (0.94, 3.45) | 0.108 | 0.85<br>(-0.10, 1.81) |
|  | 101-500 m within strata of household food animals | 1.10 (0.76, 1.60) | 1.15 (0.73, 1.83) | - | - |
|  | ≤ 100 m within strata of household food animals | 0.85 (0.46, 1.57) | 1.24 (0.82, 1.86) | - | - |

Log-binomial regression models with generalized estimating equations included interaction terms between commercial and household food animal exposure variables, and included the following covariates: caregiver education, asset score, child age and sex, and child antibiotic use in the last 3 months. N=1,940 observations across 1,677 child fecal samples (including 910 total 3GCR-EC isolates) for 594 children. CI: 95% confidence interval. 3GCR-EC: third-generation cephalosporin-resistant *E. coli*. RERI: relative excess risk due to interaction.

Additional risk factors of ESBL-EC carriage among children in our study site included placing animal feces on household land/crops (RR: 1.63; 95% CI: 1.09, 2.46), household dog ownership (RR: 1.35; 95% CI: 1.00, 1.83), and child pet contact in the last 3 months (RR: 1.54; 95% CI: 1.10, 2.16) (Figure 4, Table S10). Potential risk factors that were marginally associated with ESBL-EC carriage among children included the presence of ESBL-EC+ animal feces in the yard (RR: 1.39; 95% CI: 0.96, 2.02) and household pig ownership (RR: 1.46; 95% CI: 0.94, 2.28) (Figure 4, Table S10).

## Discussion

This study of seven semi-rural parishes in Ecuador leveraged household survey data, satellite imagery, and geographic information systems to assess individual and combined exposures to household and commercial food animal production, as well as other risk factors related to domestic animals and hygiene behavior, and the associated risk of antibiotic-resistant *E. coli* carriage in young children. Increased density of commercial food animal production facilities, household food animal ownership, child pet contact, and rarely/never washing hands after contact with animals were risk factors for 3GCR-EC carriage. The combination of owning household food animals and living within 100 m of a drainage flow path from a commercial food animal operation may have increased the risk of ESBL-EC carriage among children. Other risk factors for ESBL-EC carriage were household dog ownership, child pet contact, and placing animal feces on household land/crops. Clinically relevant STs such as ST 10, ST 131, ST 38, and ST 117 associated with extraintestinal pathogenic *E. coli* (ExPEC) were detected in child fecal samples, highlighting the public health significance of community-acquired ESBL-producing *E. coli* carriage (55). The results of this study emphasize the need for a One Health approach in the control and prevention of antibiotic-resistant bacterial infections. This is especially important as commercial food animal production expands within emerging markets.

Epidemiologic studies have established a clear link between commercial food animal production and community-acquired antibiotic-resistant infections among commercial animal farm workers, their household contacts, and community members (18,56–60). Epidemiological research on the link between small-scale food animal production and community-acquired resistance in humans, however, has been limited. Previous studies have used samples that are not spatiotemporally matched with household-level exposures, cross-sectional study designs, small sample sizes, or descriptive statistics, only (61–64). These methodological limitations prevent the reliable estimation of associations between small-scale food animal exposures and community-acquired antibiotic-resistant infections. This study attempted to address these gaps by leveraging repeated measures data from 600 households in which household-level exposure data are spatiotemporally matched with outcome data. With our robust longitudinal One Health study design, we were able to estimate the impacts of food animal exposures, domestic animal exposures, and hygiene practices on antibiotic-resistant *E. coli* carriage in children.

Research in this same study site previously provided evidence for both horizontal gene transfer and clonal spread of 3GCR-EC in children and animals within and between households in the study site (47,65). Animal waste management and handling practices were poor in a majority of households with clonal relationships between 3GCR-EC in children and animals (47). Though evidence of resistant bacteria transmission between backyard chickens, dogs, and humans has been documented in this study site and study period through previous analyses (47,65), it is unclear whether antibiotic use in household food animals and domestic animals or human antibiotic use is driving selection of resistant bacteria in these communities. In fact, reported antibiotic use in children and household animals was low in this study site and participants had limited knowledge about antibiotics and antibiotic stewardship (33). A recent qualitative study in the same communities found that small-scale poultry and livestock producers typically rely on low-cost traditional veterinary practices rather than administering antibiotics (66). Notably, antibiotic use in children and domestic animals declined throughout the study period. One possible explanation is that public health measures such as the SARS-CoV2 pandemic lockdowns may have curbed infectious disease transmission in humans (and indirectly, in animals), reducing the need for antibiotic treatment (67). This hypothesis is supported by the apparent reduction in reported child illness from cycle three (before pandemic lockdowns) to cycle five (after lockdowns began) in our study. These observed secular trends appear to be nondifferential by household food animal ownership (Table S11).

While antibiotic use in household food animals may not be a primary driver of resistance in this area, commercial-scale production operations administering high volumes of antibiotics may be largely responsible for driving the emergence and selection of resistant bacteria (1,18). Results from this study suggest that household food animals and domestic animals still play an important role in determining risk of antibiotic-resistant *E. coli* carriage. Household animals may act as a vector for transmission of resistant bacteria between environmental reservoirs contaminated by commercial food animal waste and humans with whom they come into contact. GPS-tracked movement patterns of free-range poultry in northwestern Ecuador confirmed that backyard chickens (not given antibiotics) travel an average of 17 m from their household, and that this range overlapped with small-scale farms of broiler chickens (given antibiotics) (68). Free-roaming dogs are also common in urban and rural Quito (69), and have been shown to roam up to 28 km from their homes on average in rural Southern Chile (70). While some backyard chickens in our study site were kept in coops, some free-range chickens and free-roaming dogs may have been exposed to environmental contamination from nearby commercial operations. Exposed domestic animals may be carriers of 3GCR-EC and ESBL-EC and could increase the risk of ARB transmission to household members through contact with companion animals, food animals, or their feces. In fact, children with recent contact with pets, from households with ESBL+ animal feces in the yard, or from households that applied animal waste to household food crops had a greater risk of ESBL-EC colonization in our study.

A significant challenge in identifying the source of ARB remains: there is a lack of available data on antibiotic usage and resistance in intensive, commercial food animal operations and their effluent in Ecuador due to limited oversight and surveillance. To accurately characterize the extent to which antibiotic use in commercial production drives community-acquired antibiotic-resistant infections, policies should require that commercial food animal operations monitor and report antibiotic use and conduct routine surveillance for antibiotic-resistant bacteria in food animals, food animal production waste, nearby environmental reservoirs, and food animal products. Future studies should attempt to collect this data to characterize ARB in commercial food animal production settings in Quito. Another limitation of the present study is the change in the number of *E. coli* isolated per sample midway through the study. We attempted to address this by analyzing data at the isolate level rather than the sample level, adjusting for imbalanced data in our statistical approach, and including a sensitivity analysis using only the first isolate per sample. Despite some slight differences in point estimates, the overall findings were robust to sensitivity analyses, suggesting a low risk of bias in our outcome assessment methods. Another limitation is that we only isolated 3GCR-EC in order to improve detection of ESBL-EC, limiting our findings to this specific type of antibiotic-resistant bacterial species. We did not identify other ESBL-producing enterobacteriaceae, such as *Klebsiella pneumoniae*, which also have clinical importance given the high mortality rates associated with ESBL-producing *K. pneumoniae* infections (71). Finally, this study was halted in the middle of the fourth cycle of data collection due to SARS-CoV2 lockdowns in March 2020 and did not resume until April 2021, resulting in significant participant dropout. This loss to follow-up may have induced some selection bias, though we aimed to address the imbalanced nature of the data in our statistical approach using non-parametric methods.

Prevalence estimates in the literature suggest that community-acquired ESBL-EC carriage in healthy populations is increasing globally, with a recent pooled estimate of 21% in 2015-2018 (15). However, there are few studies estimating community-acquired ESBL-producing infections in South America, and estimates are variable. Bezabih et al. (2021) estimated a pooled ESBL-EC prevalence of <10% for the Americas, while a 2015 review of ESBL-E estimated a prevalence of 2% for the Americas (72); both reviews included estimates from studies from the United States in pooled estimates and did not include any studies from Ecuador. Of note, studies in these reviews used selective media to screen for 3GCR-EC or ESBL-EC prior to ESBL confirmatory testing, comparable to the methods used in the present study. A 2008 multi-country study in South and Central America (not including Ecuador) detected ESBL-producing bacteria in 31% of community-acquired intra-abdominal infections (73). The high prevalence of 3GCR-EC, ESBL-EC, and MDR *E. coli* carriage among healthy children in our study site is concerning. Over 60% of children were carriers of 3GCR-EC with frequent detection of *bla*_CTX-M_ genes and 25% were carriers of ESBL-EC at least once throughout the study period, with 86% of all 3GCR-EC being MDR. Even among non-ESBL-producing *E. coli* isolates based on phenotypic testing, *bla* genes encoding for extended-spectrum beta-lactam resistance were detected in over 75% of 3GCR-EC isolates. CTX-M-encoding genes *bla*_CTX-M-55_, *bla*_CTX-M-65_, and *bla*_CTX-M-15_ were the most frequently detected *bla* genes in this food animal producing region of Ecuador. *bla*_CTX-M-55_, *bla*_CTX-M-65_ and *bla*_CTX-M-15_ are dominant in food animals such as chickens, pigs, and cattle, as well as meat products in China, South Korea, Hong Kong, Canada, Portugal, and elsewhere (74–78). The distribution of CTX-M-encoding genes in our study site suggests that food animal production plays a critical role in driving the community spread of 3GCR-EC and ESBL-EC in this region of Ecuador.

Evidence from this study supports several policy recommendations around antibiotic use and waste management at food animal production facilities. Local and national policies requiring improved waste management practices in large-scale commercial food animal production operations are needed in LMICs (79). Strategies may include containing and/or diverting animal waste, moving animal grazing areas away from waterways, transporting excess waste to more remote areas with sufficient non-food crop land to apply waste as fertilizer, and monitoring levels of nutrients and antibiotic-resistant bacteria in soil and waterways near discharge points. Governments could implement a permitting process that requires commercial operations to submit a waste management plan for approval by agriculture and environmental protection agencies. For example, in the United States, the Environmental Protection Agency established the National Pollution Discharge Elimination System, which regulates the discharge of pollution from point sources at large animal feeding operations and other industrial sites to bodies of water. Agencies could provide subsidies to support producers with initial costs of improving waste management. Policies and regulations should be tailored to farm size; some practices may be cost-prohibitive for small-scale producers, so appropriate financial incentives should be used to promote best management practices among small food animal farms (80). Finally, national policies should follow global recommendations to restrict the use of clinically important antibiotics like third-generation cephalosporins in food animals (81). Restricting antibiotic use in cattle farms has been shown to reduce detection of *bla*_CTX-M_ genes in cattle (82). Use of third-generation cephalosporins in food animal production should be limited to disease treatment purposes only, and should require veterinarian prescriptions in order to purchase. To improve the effectiveness of such policies, antibiotic stewardship training programs from a One Health lens could be offered to physicians, veterinarians, and food animal producers in Ecuador with an emphasis on the growing antibiotic resistance crisis.

Our study underscores the need for increased monitoring of waste management practices and improved surveillance of antibiotic use and community-acquired antibiotic resistance in LMICs with widespread food animal production. Increased contact with domestic animals, household food animal ownership, and proximity to large-scale food animal production operations – especially in high-density areas – increased the risk of antibiotic-resistant bacteria carriage in young children. With zoonotic infectious disease risks and hygiene-related prevention strategies at the forefront of public health messaging due to the SARS-CoV2 pandemic, national governments should prioritize policies and communication strategies that promote improved food animal waste management and safe hygiene practices to reduce the prevalence of antibiotic-resistant infections.

## Data Availability

Raw reads from isolates sequenced in this study are available at the NCBI Short Read Archive(SRA) under BioProject accession no. PRJNA861272. All data used to perform analyses in the present study are currently available upon reasonable request to the authors.

## Acknowledgements

We sincerely thank the data collection team in Ecuador for their hard work, time, and dedication to this research, especially through the SARS-CoV2 pandemic. Thanks to Professor Lisa Barcellos, Professor Ayesha Mahmud, and Professor Ellen Eisen for their invaluable input on the methodological approach in the early phases of this analysis. Finally, we are extremely grateful for the generosity and commitment of our study participants in Quito, without whom this work would not be possible.

## Declarations

### Data Availability

Raw reads from isolates sequenced in this study are available at the NCBI Short Read Archive (SRA) under BioProject accession no. PRJNA861272.

### Human & Animal Ethics Statement

The study was approved by the Office for Protection of Human Subjects (OPHS) at the University of California, Berkeley (IRB# 2019-02-11803) and by the Bioethics Committee at the Universidad San Francisco de Quito (#2017-178M), and the Ecuadorian Health Ministry (#MSPCURI000243-3).

