## Supplemental Information for "Risk factors for extended-spectrum beta-lactamase (ESBL) producing *E. coli* carriage among children in a food animal producing region of Quito, Ecuador"

### Supplemental Materials

**Table S1.** Prevalence of third-generation cephalosporin-resistant, extended-spectrum beta-lactamase, multidrug-resistant, and extensively drug-resistant *E. coli* among children.

| | **Data Collection Cycle *n* (%)** | | | | | |
|---|---|---|---|---|---|---|
| | 1 | 2 | 3 | 4 | 5 | Total |
| Total child fecal samples | 376 (100) | 361 (100) | 371 (100) | 230 (100) | 361 (100) | 1699 (100) |
| *ESBL-EC+* samples | 44 (12) | 48 (13) | 26 (7) | 37 (16) | 28 (8) | 183 (11) |
| *MDR-EC+* samples | 164 (44) | 151 (42) | 130 (35) | 76 (33) | 53 (15) | 574 (34) |
| *XDR-EC+* samples | 89 (24) | 78 (22) | 63 (17) | 27 (12) | 15 (4) | 272 (16) |
| Total 3GCR-EC isolates | 273 (100) | 234 (100) | 217 (100) | 115 (100) | 71 (100) | 910 (100) |
| *ESBL-EC isolates* | 51 (19) | 52 (22) | 29 (13) | 40 (35) | 28 (39) | 200 (22) |
| *MDR-EC isolates* | 234 (86) | 219 (94) | 188 (87) | 93 (81) | 53 (75) | 787 (86) |
| *XDR-EC isolates* | 118 (43) | 97 (41) | 76 (35) | 29 (25) | 15 (21) | 335 (37) |

3GCR-EC: third-generation cephalosporin-resistant *E. coli*. ESBL-EC: extended-spectrum beta-lactamase producing *E. coli* based on phenotypic testing. MDR-EC: multidrug-resistant (phenotypically resistant to 3 or more drug classes including third-generation cephalosporin) *E. coli*. XDR-EC: extensively drug-resistant (phenotypically resistant to 5 or more drug classes including third-generation cephalosporin) *E. coli*.

**Table S2.** Proportion of third-generation cephalosporin-resistant *E. coli* (3GCR-EC) isolates resistant to individual antibiotics in phenotypic susceptibility testing by data collection cycle.

| Cycle | 3GCR-EC Isolates | **3GCR-EC Isolates Resistant to Antibiotic *n* (%)** | | | | | | | | | |
|---|---|---|---|---|---|---|---|---|---|---|---|
| | | AM | CAZ | CIP | CTX | CZ | FEP | GM | IPM | SXT | TE |
| 1 | 273 (100) | 271 (99) | 71 (26) | 136 (50) | 252 (92) | 273 (100) | 119 (44) | 46 (17) | 2 (1) | 182 (67) | 201 (74) |
| 2 | 234 (100) | 233 (100) | 57 (24) | 118 (50) | 222 (95) | 233 (100) | 104 (44) | 48 (21) | 1 (0) | 153 (65) | 182 (78) |
| 3 | 217 (100) | 215 (99) | 48 (22) | 100 (46) | 199 (92) | 215 (99) | 54 (25) | 32 (15) | 1 (0) | 144 (66) | 152 (70) |
| 4 | 115 (100) | 118 (100) | 28 (24) | 42 (36) | 107 (91) | 118 (100) | 33 (28) | 16 (14) | 0 (0) | 61 (52) | 78 (66) |
| 5 | 71 (100) | 67 (94) | 11 (15) | 21 (30) | 58 (82) | 68 (96) | 20 (28) | 8 (11) | 1 (1) | 39 (55) | 40 (56) |
| Total | 910 (100) | 904 (99) | 215 (24) | 417 (46) | 838 (92) | 907 (100) | 330 (36) | 150 (16) | 5 (1) | 579 (64) | 653 (72) |

3GCR-EC: third-generation cephalosporin-resistant *E. coli;* AM: ampicillin; CAZ: ceftazidime; CIP: ciprofloxacin; CTX: cefotaxime; CZ: cefazolin; FEP: cefepime; GM: gentamicin; IPM: imipenem; SXT: trimethoprim/sulfamethoxazole; TE: tetracycline.

**Table S3.** Antibiotic resistance of 3GCR-EC isolates from animal fecal samples (one colony isolated per fecal sample) collected at the same households as child fecal samples, stratified by animal species.

| Species | H | S | 3GCR-EC Isolates | **3GCR-EC Isolates Resistant to Antibiotics *n* (%)** | | | | | | | | | | |
|---|---|---|---|---|---|---|---|---|---|---|---|---|---|---|
| | | | | ESBL | AM | CAZ | CIP | CTX | CZ | FEP | GM | IPM | SXT | TE |
| Chickens | 181 | 364 | 255 (100) | 53 (21) | 255 (100) | 65 (26) | 109 (43) | 247 (97) | 254 (99) | 107 (42) | 44 (17) | 1 (<1) | 161 (63) | 205 (80) |
| Dogs | 354 | 903 | 621 (100) | 145 (23) | 616 (99) | 145 (23) | 307 (49) | 598 (96) | 619 (99) | 245 (39) | 128 (21) | 1 (<1) | 417 (67) | 503 (81) |
| Pigs | 55 | 366 | 80 (100) | 22 (28) | 80 (100) | 29 (36) | 34 (43) | 75 (94) | 80 (100) | 35 (44) | 17 (21) | 0 (0) | 47 (59) | 62 (78) |
| Waterfowl/ pheasants* | 81 | 101 | 67 (100) | 15 (22) | 67 (100) | 15 (22) | 14 (21) | 67 (100) | 67 (100) | 32 (48) | 11 (16) | 0 (0) | 39 (58) | 61 (91) |
| Other** | 127 | 138 | 38 (100) | 5 (13) | 38 (100) | 7 (18) | 7 (18) | 37 (97) | 38 (100) | 15 (39) | 6 (16) | 0 (0) | 20 (53) | 24 (63) |
| Total | 376 | 1,871 | 1,191 (100) | 240 (20) | 1,056 (89) | 261 (22) | 471 (40) | 1,024 (86) | 1,058 (89) | 434 (36) | 206 (17) | 2 (<1) | 684 (57) | 855 (72) |

*Waterfowl/pheasants include ducks, geese, and quail. **Other includes cows, goats, sheep, horses, llamas, Guinea pigs, cats, rabbits, and other birds. HH: households; S: stool samples; 3GCR-EC: third-generation cephalosporin-resistant *E. coli*; ESBL: extended-spectrum beta-lactamase producing; AM: ampicillin; CAZ: ceftazidime; CIP: ciprofloxacin; CTX: cefotaxime; CZ: cefazolin; FEP: cefepime; GM: gentamicin; IPM: imipenem; SXT: trimethoprim/sulfamethoxazole; TE: tetracycline.

**Table S4.** Prevalence of clinically important sequence types (ST) among sequenced 3GCR-EC isolates (N=571) from child fecal samples.

| ST | No. Isolates (%) | |
|---|---|---|
| 10 | 41 | (7.18) |
| 38 | 27 | (4.73) |
| 354 | 23 | (4.03) |
| 131 | 20 | (3.50) |
| 117 | 19 | (3.33) |
| 69 | 9 | (1.58) |
| 23 | 7 | (1.23) |
| 617 | 7 | (1.23) |
| 90 | 7 | (1.23) |
| 58 | 6 | (1.05) |
| 224 | 5 | (0.88) |
| 1193 | 4 | (0.70) |
| 1196 | 3 | (0.53) |
| 457 | 3 | (0.53) |
| 167 | 2 | (0.35) |
| 410 | 1 | (0.18) |
| 88 | 1 | (0.18) |
| 95 | 1 | (0.18) |

**Table S5.** Proportion of 3GCR-EC isolates with beta-lactamase genes (among 15 most prevalent) detected in whole-genome sequences, stratified by phenotypic ESBL production.

| | ESBL- (n=426) | | | ESBL+ (n=145) | | |
|---|---|---|---|---|---|---|
| Rank | Gene | *n* | *(%)* | Gene | *n* | *(%)* |
| 1 | $bla_{TEM-141}$ | 100 | (23.47) | $bla_{CTX-M-55}$ | 47 | (32.41) |
| 2 | $bla_{CTX-M-55}$ | 83 | (19.48) | $bla_{TEM-141}$ | 31 | (21.38) |
| 3 | $bla_{TEM-1B}$ | 72 | (16.9) | $bla_{TEM-1B}$ | 23 | (15.86) |
| 4 | $bla_{CTX-M-65}$ | 50 | (11.74) | $bla_{CTX-M-15}$ | 21 | (14.48) |
| 5 | $bla_{CTX-M-15}$ | 49 | (11.5) | $bla_{OXA-1}$ | 13 | (8.97) |
| 6 | $bla_{TEM-104}$ | 44 | (10.33) | $bla_{TEM-102}$ | 11 | (7.59) |
| 7 | $bla_{CMY-2}$ | 41 | (9.62) | $bla_{CTX-M-65}$ | 10 | (6.9) |
| 8 | $bla_{TEM-102}$ | 36 | (8.45) | $bla_{TEM-105}$ | 10 | (6.9) |
| 9 | $bla_{CTX-M-3}$ | 31 | (7.28) | $bla_{SHV-12}$ | 9 | (6.21) |
| 10 | $bla_{TEM-105}$ | 29 | (6.81) | $bla_{TEM-104}$ | 9 | (6.21) |
| 11 | $bla_{CTX-M-27}$ | 17 | (3.99) | $bla_{CTX-M-101}$ | 7 | (4.83) |
| 12 | $bla_{CTX-M-8}$ | 12 | (2.82) | $bla_{CTX-M-27}$ | 5 | (3.45) |
| 13 | $bla_{OXA-1}$ | 11 | (2.58) | $bla_{SHV-5}$ | 4 | (2.76) |
| 14 | $bla_{TEM-176}$ | 11 | (2.58) | $bla_{TEM-1A}$ | 4 | (2.76) |
| 15 | $bla_{CTX-M-14b}$ | 10 | (2.35) | $bla_{CTX-M-14b}$ | 3 | (2.07) |

**Table S6.** Prevalence of beta-lactamase resistance genes among sequenced 3GCR-EC isolates from children, stratified by parishes and intensity of commercial food animal production.

| | | CMY | | CTX-M | | OXA | | SHV | | TEM | |
|---|---|---|---|---|---|---|---|---|---|---|---|
| Parish | CFOs* | *n* | *(%)* | *n* | *(%)* | *n* | *(%)* | *n* | *(%)* | *n* | *(%)* |
| **Low-intensity** | | | | | | | | | | | |
| Tababela (n=13) | 2 | 1 | (7.69) | 8 | (61.54) | 4 | (30.77) | 0 | (0.00) | 3 | (23.08) |
| Tumbaco (n=39) | 8 | 2 | (5.13) | 27 | (69.23) | 3 | (7.69) | 1 | (2.56) | 23 | (58.97) |
| Checa (Chilpa) (n=84) | 6 | 7 | (8.33) | 57 | (67.86) | 3 | (3.57) | 5 | (5.95) | 50 | (59.52) |
| Pifo (n=129) | 8 | 9 | (6.98) | 78 | (60.47) | 4 | (3.10) | 4 | (3.10) | 56 | (43.41) |
| **High-intensity** | | | | | | | | | | | |
| El Quinche (n=30) | 32 | 3 | (10.00) | 24 | (80.00) | 0 | (0.00) | 1 | (3.33) | 20 | (66.67) |
| Puembo (n=39) | 36 | 0 | (0.00) | 29 | (74.36) | 2 | (5.13) | 1 | (2.56) | 19 | (48.72) |
| Yaruqui (n=234) | 35 | 26 | (11.11) | 143 | (61.11) | 12 | (5.13) | 14 | (5.98) | 122 | (52.14) |

*CFOs=commercial food animal operations.

**Table S7.** Average number of total antibiotic resistance genes (ARGs) per third-generation cephalosporin-resistant *E. coli* isolate from children, stratified by parish.

| | | Total ARGs | |
|---|---|---|---|
| Parish | CFOs* | Mean | (SD) |
| **Low-intensity** | | | |
| Tababela (n=13) | 2 | 10.78 | (2.49) |
| Tumbaco (n=39) | 8 | 10.48 | (4.93) |
| Checa (Chilpa) (n=84) | 6 | 10.01 | (4.70) |
| Pifo (n=129) | 8 | 9.52 | (4.12) |
| **High-intensity** | | | |
| El Quinche (n=30) | 32 | 8.29 | (4.66) |
| Puembo (n=39) | 36 | 10.07 | (4.21) |
| Yaruqui (n=234) | 35 | 9.66 | (4.25) |

*CFOs=commercial food animal operations.

**Table S8.** Prevalence of CTX-M-type genes among sequenced third-generation cephalosporin-resistant *E. coli* isolates from children, stratified by parish and intensity of commercial food animal production.

| | **Low-intensity** | | | | | | | | **High-intensity** | | | | | |
|---|---|---|---|---|---|---|---|---|---|---|---|---|---|---|
| | Tababela (n=13) | | Tumbaco (n=39) | | Checa (Chilpa) (n=84) | | Pifo (n=129) | | El Quinche (n=30) | | Puembo (n=39) | | Yaruqui (n=234) | |
| | *n* | *%* | *n* | *%* | *n* | *%* | *n* | *%* | *n* | *%* | *n* | *%* | *n* | *%* |
| *bla*$_{CTX-M-55}$ | 2 | 15.38 | 12 | 30.77 | 18 | 21.43 | 27 | 20.93 | 4 | 13.33 | 12 | 30.77 | 55 | 23.50 |
| *bla*$_{CTX-M-65}$ | 2 | 15.38 | 3 | 7.69 | 10 | 11.90 | 13 | 10.08 | 2 | 6.67 | 4 | 10.26 | 25 | 10.68 |
| *bla*$_{CTX-M-15}$ | 4 | 30.77 | 4 | 10.26 | 8 | 9.52 | 14 | 10.85 | 13 | 43.33 | 6 | 15.38 | 21 | 8.97 |
| *bla*$_{CTX-M-27}$ | 0 | 0.00 | 3 | 7.69 | 1 | 1.19 | 5 | 3.88 | 1 | 3.33 | 1 | 2.56 | 11 | 4.70 |
| *bla*$_{CTX-M-3}$ | 0 | 0.00 | 3 | 7.69 | 9 | 10.71 | 9 | 6.98 | 0 | 0.00 | 0 | 0.00 | 11 | 4.70 |
| *bla*$_{CTX-M-14b}$ | 0 | 0.00 | 0 | 0.00 | 5 | 5.95 | 0 | 0.00 | 0 | 0.00 | 1 | 2.56 | 7 | 2.99 |
| *bla*$_{CTX-M-101}$ | 0 | 0.00 | 0 | 0.00 | 2 | 2.38 | 2 | 1.55 | 0 | 0.00 | 0 | 0.00 | 5 | 2.14 |
| *bla*$_{CTX-M-8}$ | 0 | 0.00 | 1 | 2.56 | 3 | 3.57 | 0 | 0.00 | 3 | 10.00 | 1 | 2.56 | 5 | 2.14 |
| *bla*$_{CTX-M-103}$ | 0 | 0.00 | 0 | 0.00 | 0 | 0.00 | 0 | 0.00 | 0 | 0.00 | 0 | 0.00 | 1 | 0.43 |
| *bla*$_{CTX-M-12}$ | 0 | 0.00 | 0 | 0.00 | 0 | 0.00 | 0 | 0.00 | 0 | 0.00 | 0 | 0.00 | 1 | 0.43 |
| *bla*$_{CTX-M-124}$ | 0 | 0.00 | 0 | 0.00 | 0 | 0.00 | 0 | 0.00 | 0 | 0.00 | 0 | 0.00 | 1 | 0.43 |
| *bla*$_{CTX-M-130}$ | 0 | 0.00 | 0 | 0.00 | 0 | 0.00 | 0 | 0.00 | 0 | 0.00 | 0 | 0.00 | 1 | 0.43 |
| *bla*$_{CTX-M-14}$ | 0 | 0.00 | 1 | 2.56 | 1 | 1.19 | 2 | 1.55 | 2 | 6.67 | 2 | 5.13 | 1 | 0.43 |
| *bla*$_{CTX-M-148}$ | 0 | 0.00 | 0 | 0.00 | 0 | 0.00 | 0 | 0.00 | 0 | 0.00 | 0 | 0.00 | 1 | 0.43 |
| *bla*$_{CTX-M-162}$ | 0 | 0.00 | 0 | 0.00 | 0 | 0.00 | 1 | 0.78 | 0 | 0.00 | 0 | 0.00 | 1 | 0.43 |
| *bla*$_{CTX-M-5}$ | 0 | 0.00 | 0 | 0.00 | 0 | 0.00 | 0 | 0.00 | 0 | 0.00 | 0 | 0.00 | 1 | 0.43 |
| *bla*$_{CTX-M-64}$ | 0 | 0.00 | 0 | 0.00 | 0 | 0.00 | 0 | 0.00 | 0 | 0.00 | 1 | 2.56 | 1 | 0.43 |
| *bla*$_{CTX-M-123}$ | 0 | 0.00 | 0 | 0.00 | 0 | 0.00 | 2 | 1.55 | 0 | 0.00 | 1 | 2.56 | 0 | 0.00 |
| *bla*$_{CTX-M-10}$ | 0 | 0.00 | 0 | 0.00 | 0 | 0.00 | 1 | 0.78 | 0 | 0.00 | 0 | 0.00 | 0 | 0.00 |
| *bla*$_{CTX-M-114}$ | 0 | 0.00 | 0 | 0.00 | 0 | 0.00 | 1 | 0.78 | 0 | 0.00 | 0 | 0.00 | 0 | 0.00 |
| *bla*$_{CTX-M-144}$ | 0 | 0.00 | 0 | 0.00 | 0 | 0.00 | 1 | 0.78 | 0 | 0.00 | 0 | 0.00 | 0 | 0.00 |
| *bla*$_{CTX-M-2}$ | 0 | 0.00 | 0 | 0.00 | 1 | 1.19 | 1 | 0.78 | 0 | 0.00 | 0 | 0.00 | 0 | 0.00 |
| *bla*$_{CTX-M-164}$ | 0 | 0.00 | 0 | 0.00 | 1 | 1.19 | 0 | 0.00 | 0 | 0.00 | 0 | 0.00 | 0 | 0.00 |
| *bla*$_{CTX-M-179}$ | 0 | 0.00 | 0 | 0.00 | 1 | 1.19 | 0 | 0.00 | 0 | 0.00 | 0 | 0.00 | 0 | 0.00 |

**Table S9.** Sensitivity analysis results for associations between food-animal exposures and 3GCR-EC and ESBL-EC including only one isolate per child fecal sample.

| | | **Relative Risks for 3GCR-EC (95% CI)** | | **Relative Risks for ESBL-EC (95% CI)** | |
|---|---|---|---|---|---|
| | | No household food animals | Household food animals | No household food animals | Household food animals |
| **Commercial Food Animal Operations in 5 km Radius** | ≤ 5 | 1.00 (ref) | 1.25 (0.97, 1.61) | 1.00 (ref) | 1.17 (0.65, 2.11) |
| | > 5 | 1.30 (1.10, 1.53) | 1.26 (1.04, 1.53) | 1.10 (0.74, 1.64) | 1.07 (0.67, 1.70) |
| | ≥ 5 within strata of household food animals | 1.30 (1.10, 1.53) | 1.01 (0.79, 1.29) | 1.10 (0.74, 1.64) | 0.92 (0.49, 1.70) |
| **Distance to Nearest Commercial Operation** | ≥ 1.5 km | 1.00 (ref) | 1.07 (0.86, 1.35) | 1.00 (ref) | 0.78 (0.44, 1.39) |
| | < 1.5 km | 1.14 (0.97, 1.33) | 1.21 (1.02, 1.45) | 0.90 (0.61, 1.32) | 1.13 (0.73, 1.77) |
| | < 1.5 km within strata of household food animals | 1.14 (0.97, 1.33) | 1.13 (0.90, 1.42) | 0.90 (0.61, 1.32) | 1.45 (0.79, 2.64) |
| **Distance to Commercial Operation Drainage Path** | > 500 m | 1.00 (ref) | 0.98 (0.79, 1.23) | 1.00 (ref) | 0.83 (0.45, 1.52) |
| | 101-500 m | 0.98 (0.82, 1.16) | 1.16 (0.89, 1.52) | 1.07 (0.71, 1.6) | 1.23 (0.61, 2.49) |
| | ≤ 100 m | 1.05 (0.84, 1.31) | 1.10 (0.85, 1.43) | 0.77 (0.40, 1.49) | 1.76 (0.83, 3.72) |
| | 101-500 m within strata of household food animals | 0.98 (0.82, 1.16) | 1.15 (0.92, 1.42) | 1.07 (0.71, 1.6) | 1.02 (0.61, 1.72) |
| | ≤ 100 m within strata of household food animals | 1.05 (0.84, 1.31) | 1.08 (0.87, 1.34) | 0.77 (0.40, 1.49) | 1.07 (0.67, 1.70) |

Log-binomial regression models with generalized estimating equations included interaction terms between commercial and household food animal exposure variables, and included the following covariates: caregiver education, asset score, child age and sex, and child antibiotic use in the last 3 months. N=1,677 observations across 1,677 child fecal samples (including 663 total 3GCR-EC isolates) for 594 children (number of observations includes multiple isolates per fecal sample). CI: 95% confidence interval. 3GCR-EC: third-generation cephalosporin-resistant *E. coli.*

**Table S10.** Adjusted relative risks (RR), 95% confidence intervals (CI), and *P*-values for individual risk factors (see Figure 3) for 3GCR-EC and ESBL-EC carriage among children.

| | | 3GCR-EC | | ESBL-EC | |
|---|---|---|---|---|---|
| Risk Factor | N | RR (95% CI) | *P*-value | RR (95% CI) | *P*-value |
| Household animals received antibiotics in last 6 months (vs. not) | 1922 | 1.148 (0.946, 1.393) | 0.1626 | 1.149 (0.626, 2.107) | 0.6546 |
| Livestock/poultry drank irrigation water in last 3 weeks (vs. not) | 955 | 1.209 (0.94, 1.557) | 0.1397 | 1.214 (0.722, 2.043) | 0.4643 |
| Animal feces left in yard (vs. placed in trash) | 1133 | 0.883 (0.73, 1.068) | 0.2007 | 0.685 (0.403, 1.164) | 0.1616 |
| Animal feces placed on land/crops (vs. placed in trash) | 1133 | 1.042 (0.86, 1.262) | 0.6759 | 1.632 (1.086, 2.455) | 0.0186 |
| Household owns dogs (vs. none) | 1949 | 1.017 (0.9, 1.151) | 0.7826 | 1.352 (0.996, 1.834) | 0.0532 |
| Household owns chickens (vs. none) | 1949 | 1.104 (0.98, 1.243) | 0.1044 | 1.164 (0.84, 1.614) | 0.3608 |
| Household owns pigs (vs. none) | 1948 | 1.227 (1.021, 1.475) | 0.0295 | 1.458 (0.934, 2.277) | 0.0971 |
| 3GCR-EC+ animal feces in yard (vs. ESBL-EC- feces) | 1144 | 1.032 (0.897, 1.188) | 0.6573 | 1.095 (0.797, 1.505) | 0.5753 |
| ESBL-EC+ animal feces in yard (vs. 3GCR-EC- feces) | 1144 | 1.112 (0.949, 1.303) | 0.1879 | 1.392 (0.959, 2.022) | 0.0819 |
| Caregiver worked with animals in last 6 months* (vs. not) | 1949 | 0.949 (0.821, 1.098) | 0.4851 | 1.133 (0.805, 1.596) | 0.4738 |
| Child contact with livestock in last 3 months (vs. not) | 1949 | 1.038 (0.918, 1.174) | 0.5520 | 1.023 (0.76, 1.377) | 0.8801 |
| Child contact with pets in last 3 months (vs. not) | 1949 | 1.229 (1.089, 1.388) | 0.0009 | 1.542 (1.102, 2.158) | 0.0116 |
| Child played near animal feces in last 3 weeks (vs. not) | 1942 | 0.993 (0.879, 1.123) | 0.9167 | 1.09 (0.811, 1.465) | 0.5680 |
| Child rarely/never handwashes after contact with animals (vs. sometimes/always) | 1882 | 1.147 (0.98, 1.342) | 0.0872 | 1.138 (0.729, 1.775) | 0.5704 |

* including working with live animals, animal feces, or meat processing. Log-binomial regression models with generalized estimating equations included the following covariates: caregiver education, asset score, child age and sex, and child antibiotic use in the last 3 months. 3GCR-EC: third-generation cephalosporin-resistant *E. coli*; ESBL-EC: extended-spectrum beta-lactamase *E. coli*; RR: relative risk.

**Table S11.** Secular trends in caregiver-reported child illness and antibiotic use stratified by household food animal ownership.

| | Data Collection Cycle | | | | |
|---|---|---|---|---|---|
| | 1<br>*n* (%) | 2<br>*n* (%) | 3<br>*n* (%) | 4<br>*n* (%) | 5<br>*n* (%) |
| **No household food animals** | | | | | |
| *Child had diarrhea in last 7 days* | 48 (18.6) | 46 (19.2) | 28 (10.9) | 25 (15.4) | 11 (4.2) |
| Missing | 1 (0.4) | 1 (0.4) | 0 (0.0) | 0 (0.0) | 2 (0.8) |
| *Child treated for infection in last 3 months* | 77 (29.8) | 69 (28.7) | 66 (25.6) | 42 (25.9) | 21 (8.0) |
| Missing | 2 (0.8) | 1 (0.4) | 0 (0.0) | 0 (0.0) | 0 (0.0) |
| *Child took antibiotics in last 3 months* | 62 (24.0) | 46 (19.2) | 48 (18.6) | 18 (11.1) | 15 (5.7) |
| Missing | 0 (0.0) | 1 (0.4) | 0 (0.0) | 0 (0.0) | 0 (0.0) |
| **Household food animals** | | | | | |
| *Child had diarrhea in last 7 days* | 27 (24.5) | 31 (26.5) | 15 (14.0) | 11 (17.5) | 4 (4.3) |
| Missing | 1 (0.9) | 0 (0.0) | 1 (0.9) | 0 (0.0) | 0 (0.0) |
| *Child treated for infection in last 3 months* | 38 (34.5) | 29 (24.8) | 18 (16.8) | 12 (19.0) | 8 (8.7) |
| Missing | 1 (0.9) | 0 (0.0) | 0 (0.0) | 0 (0.0) | 0 (0.0) |
| *Child took antibiotics in last 3 months* | 35 (31.8) | 21 (17.9) | 15 (14.0) | 8 (12.7) | 6 (6.5) |
| Missing | 0 (0.0) | 1 (0.9) | 0 (0.0) | 0 (0.0) | 0 (0.0) |

**Figure S1.** Directed acyclic graph of causal relationship between exposures to commercial and household food animal production and ESBL-*E. coli* carriage in children.

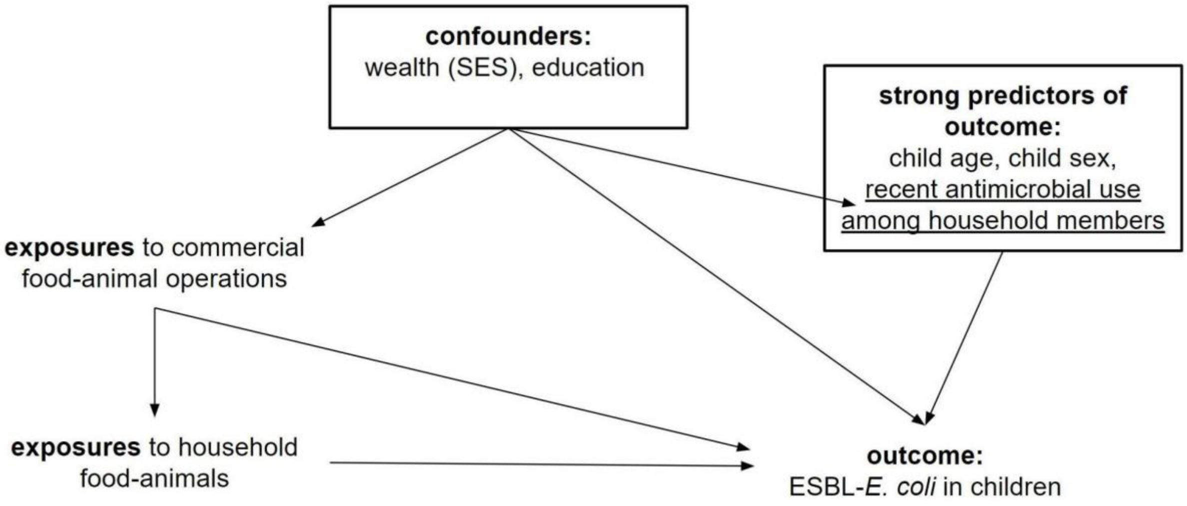

**Figure S2.** Flow chart of enrollment and follow-up by data collection cycle. Households were included in the final analysis if they had the necessary exposure, outcome, and covariate data.

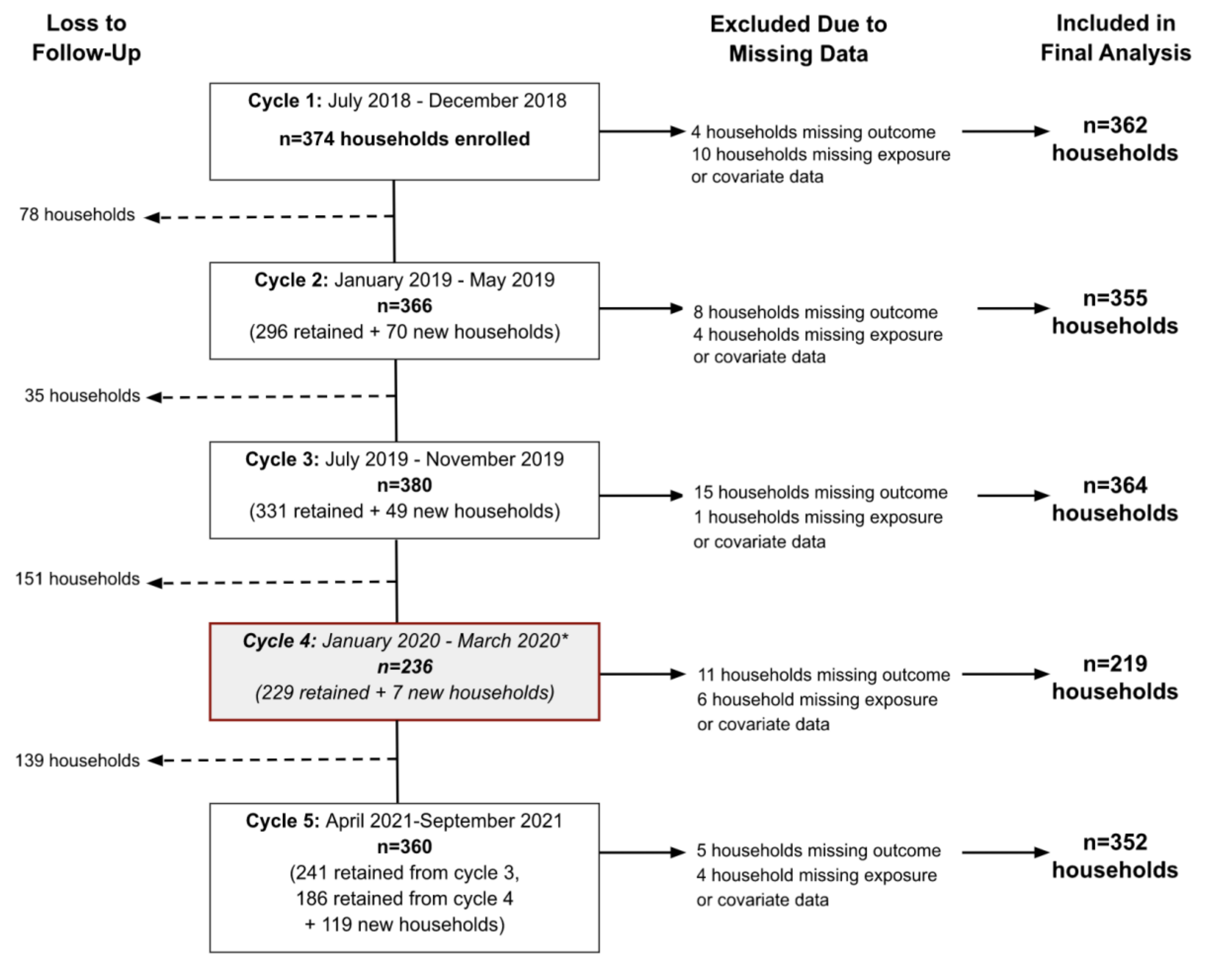

**Figure S3.** Prevalence of beta-lactamase genes (top 15 most prevalent) among sequenced third-generation cephalosporin-resistant *E. coli* isolates from children, stratified by phenotypic ESBL production.

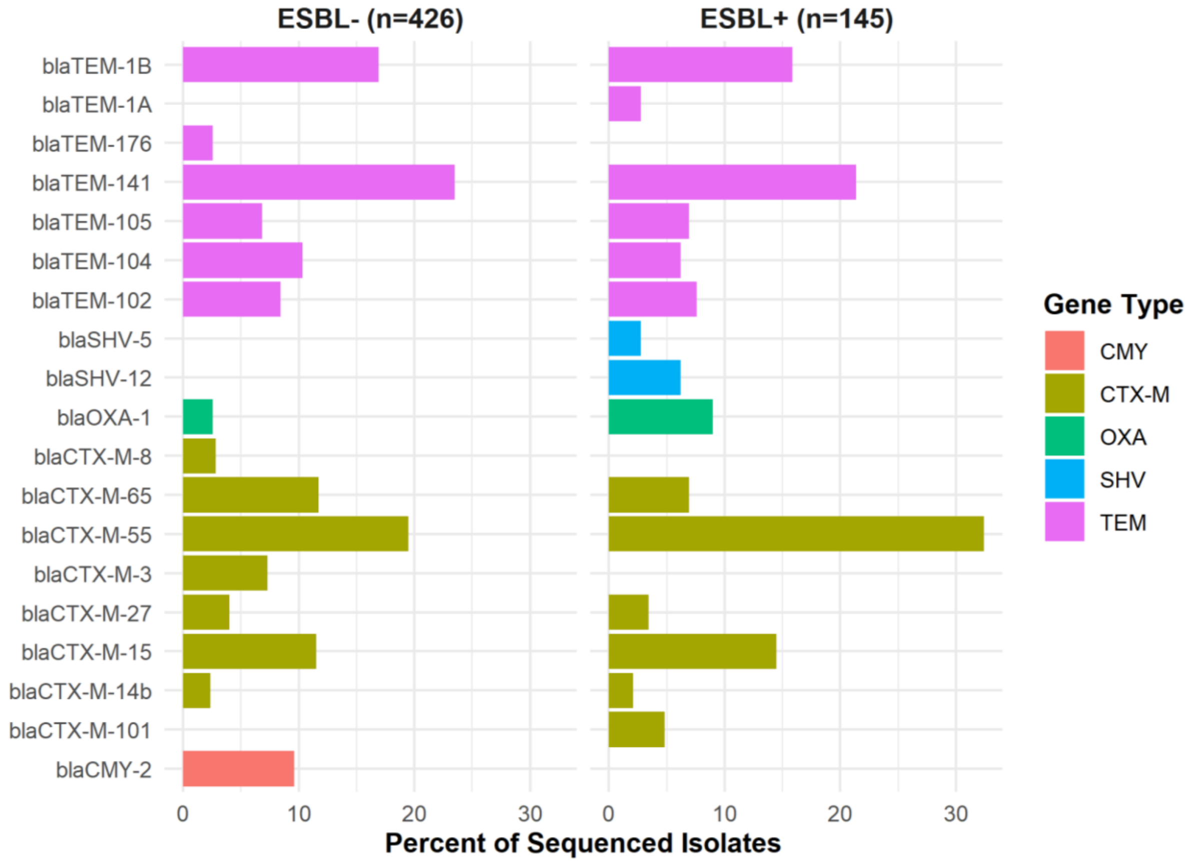

**Figure S4.** Prevalence of beta-lactamase genes by type among third-generation cephalosporin-resistant *E. coli* isolated from children, stratified by parish.

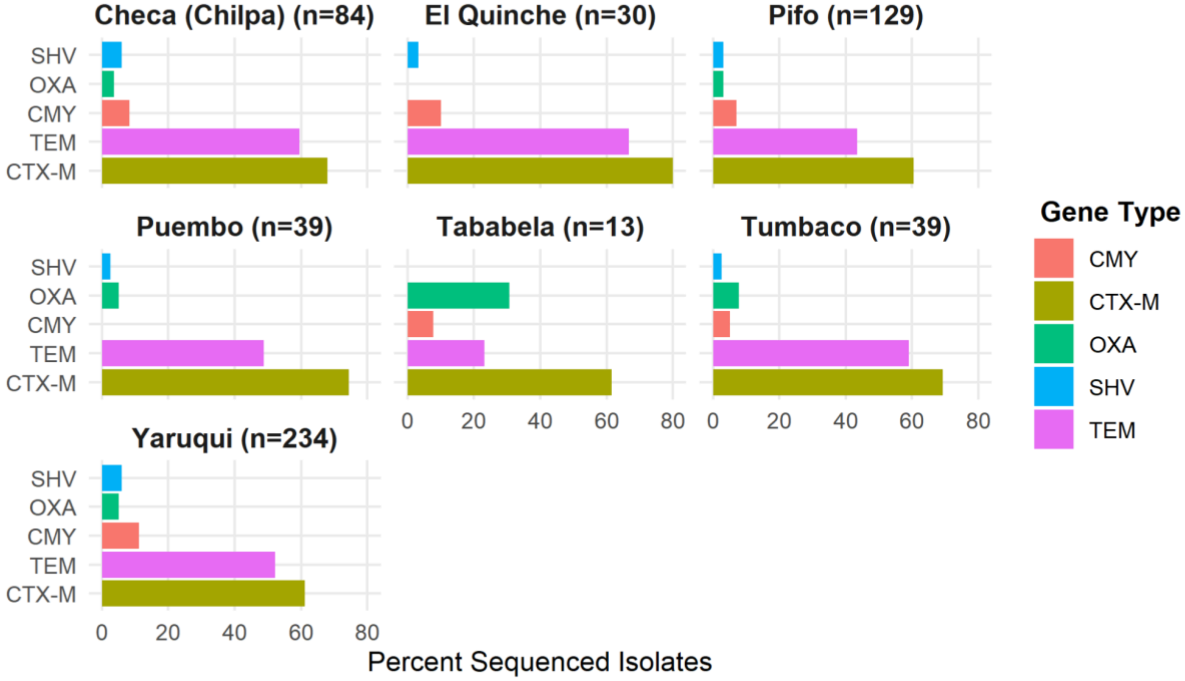
